# Genome-Wide Association Study of Obsessive-Compulsive Symptoms including 33 943 individuals from the general population

**DOI:** 10.1101/2022.11.30.22282898

**Authors:** Nora I. Strom, Christie L. Burton, Conrad Iyegbe, Talisa Silzer, Lilit Antonyan, René Pool, Mathieu Lemire, James J. Crowley, Jouke-Jan Hottenga, Volen Z. Ivanov, Henrik Larsson, Paul Lichtenstein, Patrik Magnusson, Christian Rück, Russell Schachar, Hei Man Wu, Danielle Cath, Jennifer Crosbie, David Mataix-Cols, Dorret I. Boomsma, Manuel Mattheisen, Sandra M. Meier, Dirk J.A. Smit, Paul D. Arnold

## Abstract

While 1-2% of individuals meet the criteria for a clinical diagnosis of obsessive-compulsive disorder (OCD), many more (∼15-40%) experience subclinical obsessive-compulsive symptoms (OCS) during their life. To characterize the genetic underpinnings of OCS and its genetic relationship to OCD, we conducted the largest genome-wide association study (GWAS) meta-analysis of parent- or self-reported OCS to date (N = 33,943 with complete phenotypic and genome-wide data), combining the results from seven large-scale population-based cohorts from Sweden, the Netherlands, England, and Canada (including six twin cohorts and one cohort of unrelated individuals). We found no genome-wide significant associations on the SNP or gene-level, but a polygenic risk score (PRS) based on the OCD GWAS previously published by the Psychiatric Genetics Consortium (PGC-OCD) was significantly associated with OCS (P_fixed_ = 3.06 ×10^−5^). Also, one curated gene set (Mootha Gluconeogenesis) reached Bonferroni-corrected significance (N_genes_ = 28, Beta = 0.79, SE = 0.16, P_bon_ = 0.008). Expression of genes in this set is high at sites of insulin-mediated glucose disposal. Dysregulated insulin signaling in the etiology of OCS has been suggested by a previous study describing a genetic overlap of OCS with insulin signaling-related traits in children and adolescents. We report a SNP heritability of 4.1% (P = 0.0044) in the meta-analyzed GWAS, and heritability estimates based on the twin cohorts of 33% - 43%. Genetic correlation analysis showed that OCS were most strongly associated with OCD (r_G_ = 0.72, p = 0.0007) among all tested psychiatric disorders (N = 11). Of all 97 tested phenotypes, 24 showed a significant genetic correlation with OCS, and 66 traits showed concordant directions of effect with OCS and OCD. OCS have a significant polygenic contribution and share genetic risk with diagnosed OCD, supporting the hypothesis that OCD represents the extreme end of widely distributed OCS in the population.

## 1. Introduction

Obsessive-compulsive disorder (OCD) is a common and impairing disorder characterized by persistent, intrusive thoughts and/or repetitive, ritualized behaviors. OCD is a heritable condition with an estimated heritability of 47% in a twin study (1) and a heritability based on common single nucleotide polymorphisms (SNPs) of 16-28 % (2,3). However, replicated specific genetic risk factors for OCD have yet to be identified. Several disorders co-occur with OCD such as anxiety disorders, mood disorders, anorexia nervosa, tics, among others (4–6) and these disorders share genetic risk with OCD (7,8).

For many psychiatric disorders, including OCD, it is thought that their genetic risk is continuously distributed in the general population, contributing to varying levels of symptom expression (9). This is in line with the observation that obsessive-compulsive symptoms (OCS) are relatively common in the population. For example, 13 to 38 % (10) of all adult individuals experience OCS, with even higher rates in younger individuals (11,12), but only 1 to 2 % meet the criteria for a clinical diagnosis of OCD. Studies showing a shared genetic risk between OCS and diagnosed OCD support the hypothesis that clinical OCD represents the extreme end of a continuous distribution of symptoms (12,13) and that by considering sub-clinical OCS data we can increase the population available for studying (14). As clinical OCD, also OCS measured as a quantitative trait are heritable, with estimates of 30-74% from twin studies (total heritability; 15–19) and a SNP-based heritability of 7-16% from genome-wide association studies (12,13,20). Studies of quantitative OCS have also identified genome-wide significant variants. A study by den Braber et al. (20) first reported a genome-wide significant SNP (rs8100480) in *MEF2BNB* for OCS in the Netherlands Twin Register (NTR; N = 6 931), although this was not replicated in a relatively small clinical sample of patients with OCD. When OCS in the NTR sample were meta-analyzed with diagnosed OCD in the Psychiatric Genomics Consortium (PGC) sample (N =17 992), no genome-wide significant variants were identified (12). A study by Burton et al. (13) reported a genome-wide significant hit (rs7856850) in *PTPRD* for OCS in the Spit for Science sample (N = 5 018), which was also associated with OCD in a meta-analysis of independent clinical OCD and control samples (N = 11 980). Together these studies suggest that cohort- and community-based samples may be useful for identifying genetic risk for not only OCS but also OCD.

Here we present the results of the largest GWAS meta-analysis of OCS to date, combining the results from various large-scale population-based cohorts from Sweden, the Netherlands, England, and Canada that assessed OCS with a variety of questionnaires. To further characterize our GWAS results, we conducted gene-based and gene-set analyses, as well as genetic correlation analyses with 97 other traits. Further, polygenic risk score (PRS) analysis allowed us to assess the probabilistic susceptibility of OCS using the combined risk measure of variants associated with educational attainment and several psychiatric disorders that often co-occur and genetically correlate with OCD, such as depression (DEP; 15-41% comorbidity rate (21–23)), schizophrenia (SCZ; 8-26% comorbidity rate (24)), autism spectrum disorder (ASD; 17% comorbidity rate (25)), and attention-deficit hyperactivity disorder (ADHD; 6-21% comorbidity rate (21)). We further assessed the validity of using OCS in population-based samples as a proxy for clinical OCD diagnosis by comparing the OCS GWAS to the latest GWAS of OCD (2). We assessed the association of the OCD polygenic risk score (PRS) with OCS in our samples and compared the genetic correlation patterns of OCS and OCD with other traits and disorders.

## 2. Methods

### 2.1. Cohorts & obsessive-compulsive symptom measures

Individuals included in this study stem from seven different European-ancestry cohorts, including four cohorts from the Swedish Twin Registry (STR; (26–28)), namely CATSS18 (29), CATSS24 (29), STAGE, YATSS, and one each from the Netherlands Twin Register (NTR; (30)), Spit for Science (SfS; (31,32)), and TwinsUK (33). The cohorts are predominantly population-based twin cohorts, except SfS which does not include twins, with a mean age-range between 10 and 42 years (see Table 1). CATSS is a prospective, longitudinal study of all twins born in Sweden since 1992. Here, we used data measured at age 18 (CATSS18), and/or age 24 (CATSS24), selecting only one measurement timepoint per individual (preferring the measurement at age 24 over age 18 if both measurements were completed). Data from NTR (12) and SfS (13) were included in previous GWASs. See Supplementary Material for more detailed cohort descriptions.

**Table 1:** Overview of the individual cohorts. For each individual study included in the OCS meta-analysis (STRCATSS18,STR-CATSS24, STR-STAGE, STR-YATSS, NTR, SfS, TwinsUK), the table lists the questionnaire used, total sample size included (N), the number of monozygotic twin pairs (N MZ twin pairs), the number of dizygotic twin pairs (N DZ twin pairs), the number of siblings (N siblings), the number of parents (N parents), the percentage of females and males in the total N (% females (males)), and the mean age with standard deviations (Mean age ± SD). The NTR twin pairs include 14 multiplets; twins where only one twin participated were not counted as twins; other individuals included 288 spouses of twins or siblings. Note that CATSS samples were later pooled across the two Catss cohorts (CATSS15, CATSS18) for GWAS analysis, depending on the platform they were genotyped on (GSA, PsychChip).

|  | STR<br>CATSS18 | STR<br>CATSS24 | STR<br>STAGE | STR<br>YATSS | NTR | SfS | TwinsUK |
| --- | --- | --- | --- | --- | --- | --- | --- |
| **Questionnaire** | Brief<br>Obsessive-Compulsive<br>Scale<br>(BOCS) | Obsessive<br>Compulsive<br>Inventory<br>Revised<br>(OCI-R) | 7-item<br>OCS<br>instrument | Obsessive<br>Compulsive<br>Inventory<br>Revised<br>(OCI-R) | Padua<br>Inventory<br>Revised | Toronto<br>Obsessive<br>Compulsive<br>Scale<br>(TOCS) | Obsessive<br>Compulsive<br>Inventory<br>Revised<br>(OCI-R) |
| **N** | 3870 | 2225 | 7845 | 2947 | 8550 | 5171 | 3334 |
| **N MZ twin**<br>**pairs** | 292 | 194 | 561 | 276 | 1137 | - | 269 |
| **N DZ twin**<br>**pairs** | 1230 | 661 | 991 | 646 | 1722 | - | 633 |
| **N siblings** | - | - | - | - | 859 | - | - |
| **N parents** | - | - | - | - | 2608 | - | - |
| **% females** | 56% | 57% | 60% | 63% | 64% | 48% | 92% |
| **Mean age ±**<br>**SD** | 18.53 ±<br>0.33 | 23.84 ±<br>0.32 | 33.89 ±<br>7.74 | 23.93 ± 1.78 | 41.94 ±<br>15.80 | 10.87 ± 2.75 | 56.7 ± 12.6 |

Several questionnaires were used to assess OCS across the cohorts. STR-CATSS18 employed the Brief Obsessive-Compulsive Scale (BOCS; (34)), STR-STAGE used a seven-item OCS instrument (1), while STR-CATSS24, STR-YATSS, and TwinsUK employed the OCI-R (35), excluding the hoarding and neutralizing sub-scales. In NTR, the Padua Inventory Revised was used (36,37) in the form of the 12-item abbreviated and Dutch translated version (38) excluding the rumination items, leaving 9 items on checking, washing, precision, and intrusive thoughts. In SfS, parent- or self-reported obsessive-compulsive traits within the last 6 months of visiting the Ontario Science Center were assessed using 19 items from the Toronto Obsessive Compulsive Scale (TOCS), a 21-item questionnaire described elsewhere (13,39,40). Two items related to hoarding were removed. To ensure reliable and valid symptom reporting, SfS participants below 12 years of age with self-reported OCS and above 16 years of age with parent-reported OCS were excluded (see Supplementary Table S1 for cohort-specific details on OCS questionnaires)

For all cohorts, individuals with one or more missing items were excluded and items were summed and standardized into a z-score. Distributions of the raw obsessive compulsive item scores are shown in Supplementary Figures S1-S9. The distributions of the z-transformed sum scores are shown in Supplementary Figures S10-S13.

### 2.2. Genome-wide association analysis

All participants were genotyped on SNP-arrays using DNA from saliva or blood. One part of the STR-CATSS samples was genotyped on the PsychChip genotyping array (N = 5 683), another part was genotyped on the GSA genotyping array (N = 412). For the GWAS analyses STR-CATSS cohorts (CATSS18, CATSS24) were pooled over each genotyping platform (GSA, PsychChip), forming two separate CATSS datasets (STR-CATSS-GSA and STR-CATSS-PC). Each of the seven datasets (STR-CATSS-GSA, STR-CATSS-PC, STR-YATSS, STR-STAGE, NTR, SfS, and TwinsUK) underwent stringent quality-control (QC), including the removal of non-European ancestry outliers based on PCA and imputation using the HRC (STR, NTR) or the 1000G (SfS, TwinsUK) reference sets (see the Supplementary Material for more detailed information). Together, all cohorts comprised 33 943 individuals (STR: N = 16 888, NTR: N = 8 550, SfS: N = 5 171, and TwinsUK: N = 3 334) with complete phenotypic and genotypic information (see Table 1 and Supplementary Material for a more detailed description of each cohort).

We used GCTA-fastGWA (41) to perform a mixed-linear-model GWAS within each cohort separately. fastGWA controls for population stratification by principal components and for relatedness by a SNP-derived genetic relationship matrix (GRM). In STR, NTR, and TwinsUK a sparse family GRM was defined, and each GWAS analysis included the first 10 genetic principal components, sex, age, age squared and genotyping batches as covariates. In a sparse GRM, all off-diagonal values below 0.05 are set to 0 (default), thereby capturing the same proportion of phenotypic variance as by pedigree-relatedness and accounting for the close relatedness of individuals in the data. In SfS, analyses were performed on unrelated individuals. For sibling pairs, the first enrolled sibling from each family was selected for further analysis (see Table 1 for the number of siblings included). Here, GCTA-fastGWA linear regression was performed using a full-GRM and sex, age, respondent (parent vs. child reporting), genotyping array type, principal components 1-3 and projected principal components 1-3 (see supplement for details) as covariates.

For each of the seven resulting GWAS summary statistics, variants were filtered on MAF > 1%, and imputation-quality (INFO) score > 0.8. For strand ambiguous (A/T and C/G) SNPs, those with a MAF >= 0.4 were removed, while the frequencies of those with a MAF < 0.4 were compared to frequencies in the Haplotype Reference Consortium (HRC) reference (42). Same strand orientation was assumed if the frequencies matched (i.e., the minor allele was the same in both data sets). All SNPs with matching frequencies were retained while mismatched SNPs (i.e., the minor allele was different in either dataset), assumed to be reported on different strands, were flipped according to the orientation reported in the HRC reference. Following removal of poorly genotyped SNPs, all datasets were aligned to the HRC-reference.

With these harmonized datasets, we conducted an inverse variance weighted meta-analysis utilizing METAL (43), a tool included within the Rapid Imputation for COnsortias PIpeLIne (Ricopili) (44). To identify any residual population stratification or systematic technical artifacts, we inspected the genomic control factor (Lambda and Lambda1000). The genome-wide significance threshold was set to 5×10^−08^.

### 2.3. Gene-based analyses (MAGMA/FUMA)

We performed gene-analysis and gene-set analysis using Multi-marker Analysis of GenoMic Annotation (MAGMA) (45) v1.08 as implemented in Functional Mapping and Annotation of Genome-Wide Association Studies (FUMA) v1.3.7 (46). To test genetic associations at the gene level for the combined effect of SNPs in or near protein coding genes, we applied default settings (SNP-wise model for gene analysis and competitive model for gene-set analysis). Gene-based p-values were computed by mapping SNPs to their corresponding gene(s) based on their position in the genome. Positional mapping was based on ANNOVAR annotations, and the maximum distance between SNPs and genes was set to 10 kb (default). A multiple regression model was employed while accounting for linkage disequilibrium (LD) between the markers. The 1000 Genomes phase 3 reference panel (47), excluding the MHC region, was used to adjust for gene size and LD across SNPs. Using the result of the gene-based analysis (gene level p-values), competitive gene-set analysis was performed with default parameters: 15 496 gene sets were tested for association. Gene sets were obtained from MsigDB v7.0 (see www.gsea-msigdb.org for details), including ‘*Curated gene sets*’ consisting of nine data resources including KEGG, Reactome, and BioCarta, and *‘GO terms’* consisting of three categories (biological processes, cellular components, and molecular functions).

### 2.4. Heritability and cross-disorder analyses

#### 2.4.1. Heritability estimates

Heritability estimates of each cohort were extracted from the GCTA association output. GCTA uses the restricted maximum likelihood (REML) approach (48) to estimate heritability in the GRM that is supplied to correct for relatedness in the linear association test. For the twin cohorts (STR, NTR, and TwinsUK), this means that heritability was based on the sparse GRM. For SfS, with no related individuals, the heritability was based on the full GRM. For all heritability estimates, the same covariates as in the GWAS analyses were used.

We also calculated the SNP-based heritability of the OCS meta-analysis using linkage disequilibrium (LD) score regression (LDSC; (49)). LDSC bases its calculation of SNP-based heritability on the estimated slope from the regression of the SNP effect from the GWAS on the LD score.

#### 2.4.2. Cross-trait genetic correlations

With LDSC (49) we calculated genetic correlations between OCS and 97 traits, including psychiatric, substance use, cognition & socio-economic status, personality, neurological, autoimmune, cardiovascular, anthropomorphic and fertility phenotypes (see Supplementary Table S4 for a list of the source GWAS studies used). 15 of the included traits pertain to neuroticism and include the neuroticism sum score, worry- and depressive subclusters, as well as individual neuroticism items. The genetic correlation is based on the estimated slope from the regression of the product of Z-scores from the two GWASs of interest on the LD score. It represents the genetic correlation between two traits based on all polygenic effects captured by the included SNPs. Because imputation quality is correlated with LD score, and low imputation quality generally yields lower test statistics, imputation quality is a confounder for LD score regression. We therefore filtered on INFO > 0.9, if INFO was available, and MAF > 0.01. The SNPs from the European HapMap3 (50) were used as a reference. We further compared the genetic correlation patterns between OCS and OCD with all other 96 traits, to identify concordant and discordant patterns. OCD results were based on the publicly available summary statistics from the PGC (2).

#### 2.4.3. Cross-phenotype polygenic risk score analyses

To further explore the genetic relationship between OCS in each of our datasets and other (psychiatric) phenotypes, we calculated polygenic risk-scores (PRS) based on large-scale GWAS summary statistics of OCD (*N*_*Cases*_ = 2 688, *N*_*controls*_ = 7,037 ; (2)), DEP (*N*_*Cases*_ = 170 756, *N*_*controls*_ = 329 443; excluding 23andMe (51)), SCZ (*N*_*Cases*_ = 53 386, *N*_*controls*_ = 77 258; (52)), ASD (*N*_*Cases*_ = 18 381, *N*_*controls*_ = 27 969; (53)), ADHD (*N*_*Cases*_ = 19 099, *N*_*controls*_ = 34 194; (54)), and educational attainment (*N*_*Total*_ = 245 621; EA; (55)) using PRSice2 (56) and evaluated their association with OCS in our cohorts (STR-CATSS-GSA, STR-CATSS-PC, STR-STAGE, STR-YATSS, NTR, SfS, and TwinsUK). We pre-selected p-value thresholds based on the best-performing thresholds reported in the primary publications (EA: P = 1; ADHD, ASD, OCD, SCZ: P = 0.1; DEP: P = 0.5). The PRS scores were calculated as the weighted sum of the risk allele dosages.

For STR, NTR, and TwinsUK, we employed a generalized estimating equation (GEE) (57) in R to evaluate the relationship between the PRS scores and OCS scores in each cohort. The GEE analysis takes into account the resemblance within clusters, accounting for the relatedness in the datasets. Robust standard errors (sandwich-corrected) and Z-scores are reported. For SfS we applied linear regression, as conducted within the PRSice2 pipeline, to evaluate the relationship between PRS and OCS. Contributions of the PRSs were measured through comparison of the R² of the full model (including the PRS, and covariates) minus the null model (including only covariates). The same covariates that were previously included in the respective GWASs were used. We combined the PRS estimates across all target datasets using an inverse variance meta-analysis using the metagen package in R. Cochran’s Q test (58) and Higgin’s I² (59,60) were used to examine a possible heterogeneity in PRS estimates across the cohorts. Q is calculated as the weighted sum of the squared differences between individual cohort effects and the pooled effect across cohorts, with the weights being those used in the pooling method. The I² statistic describes the percentage of variation across studies that is due to heterogeneity rather than sample variation and does, unlike Q, not inherently depend on the number of measures included in the meta-analysis. We calculated a fixed effects model to evaluate the association of each PRS with OCS, regardless of observed heterogeneity. We further calculated a random effects model if there was substantial observed heterogeneity across study sites (I² > 0.5 and/or P_Q_ < 0.05).

### 2.5. Comparability of cohorts

To identify if the summary statistics from any of the included cohorts substantially deviated from the rest, we performed leave-one-out (LOO) GWAS meta-analyses and subsequently used those datasets to conduct a set of sensitivity analyses. First, we performed sign-test analyses on the top SNPs (inclusion threshold of p = 0.0001, p = 0.00001, and p = 0.000001) using the replication module of the Ricopili pipeline. Sign-tests allow for quantification of the number of genomic regions that are independent across the different p-value thresholds and identification of how many genomic regions within the replication study have the same direction of effect as the discovery. The output (in the form of a ratio) provides an estimate of the percentage of genomic regions with the same direction of effect between any two datasets. A sign-test is a binomial test with the null-hypothesis = 0.5, with a ratio > 0.5 indicating a positive sign test which (convergence), while a ratio < 0.5 indicates divergence. We conducted two sets of sign-tests, one comparing the direction of effect for each pair-wise combination of cohorts, and one comparing each LOO meta-analysis with the respective sample that was left out. While fluctuations in the sign-tests across different p-value thresholds are expected, depending on the true association of each SNP with the phenotype, we mainly aimed to assess whether a specific cohort markedly deviated from the rest.

Following the same procedure as for the cross-trait PRS analyses described above (see previous method-section on cross-trait PRS analyses for details), we conducted LOO PRS analyses to evaluate the relationship between the PRS scores of each LOO GWAS and standardized OCS scores in the left-out cohort.

Further, we conducted genetic correlation analyses between each LOO OCD meta-analysis (LOO_NTR, LOO_SfS, LOO_STR, and LOO_TwinsUK) and the same set of 97 phenotypes as described earlier to explore a possible heterogeneity in correlation patterns depending on the included OCS cohorts.

## 3. Results

### 3.1. Genome-wide association results

The final OCS GWAS meta-analysis included 6 232 765 autosomal SNPs and no significant inflation was observed (λ = 1.027, λ_1000_ = 1.001, LDSC intercept = 1.0047, see Supplementary Figure S14 for a QQ-plot). No SNP reached genome-wide significance (p < 5×10^−08^; see Figure 1 for a Miami-plot including the Manhattan-plot of the GWAS in the upper panel). The SNP with the lowest p-value was rs113538937 (p = 6.36×10^−08^) on chromosome 4 (see Supplementary Figure S15 for a regional association plot and forest plot). The region tagged by this SNP spans 207.6kb (LD r² > 0.6) and entails the genes *SH3BP2, ADD1, MFSD10, NOP14-AS1, NOP14, GRK4, HTT-AS*, and *HTT*. Another 26 independent SNPs with a p-value < 1×10^−05^ were identified (see Supplementary Table S2 for a list of association results).

**Figure 1:**
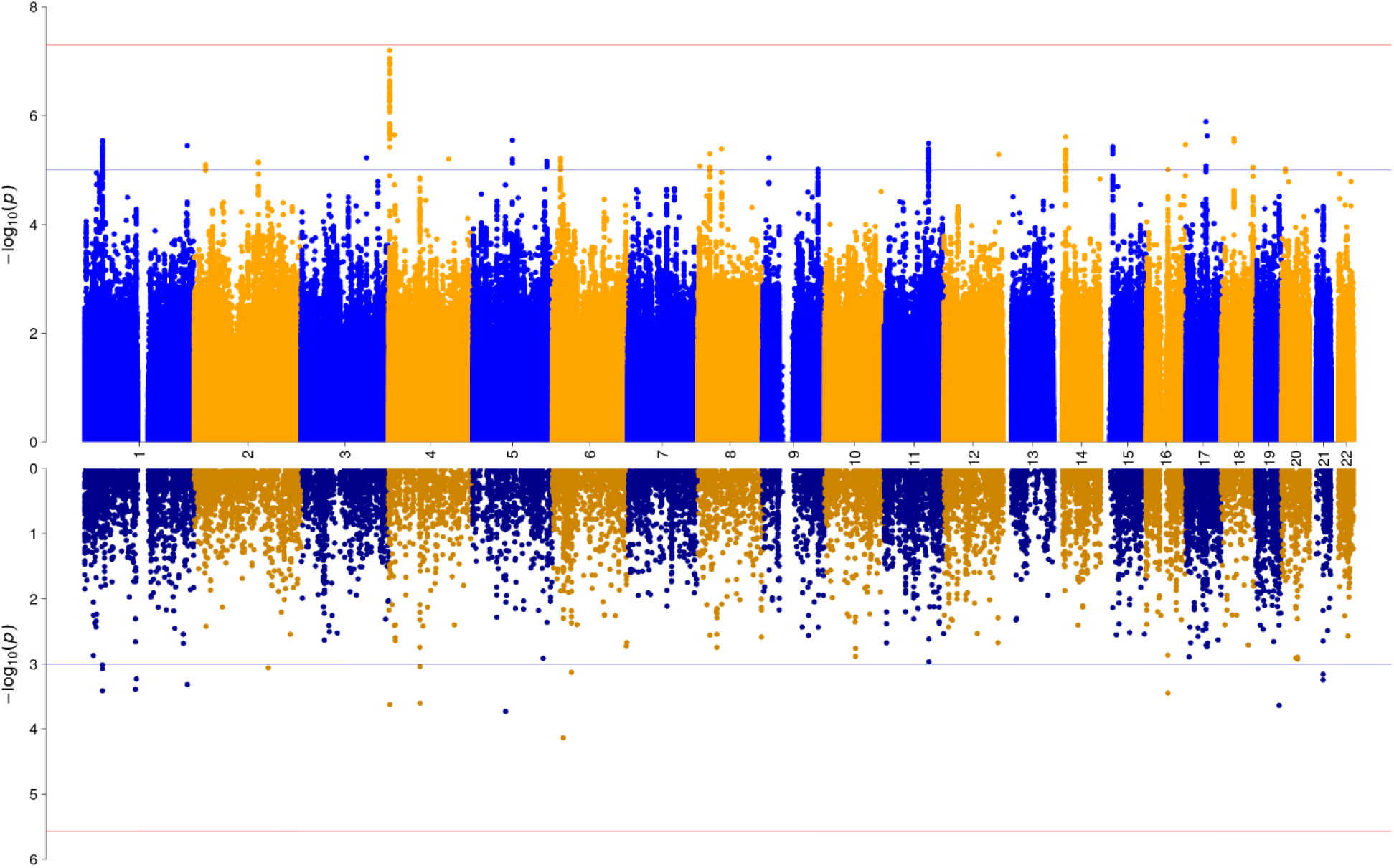
Miami plot of the association results from the GWAS meta-analysis (upper panel) and of the gene-wide association analysis (lower panel) of OCS. The y-axes represent -log10 P values for the association of SNPs/genes with OCS. The x-axis represents chromosomes 1 to 22. In the upper plot, the P-value threshold for genome-wide significance (5×10^−08^) is represented by the horizontal red line, suggestive significance (p = 1×10^−05^) by the blue line. In the lower panel, Bonferroni-corrected gene-wide significance (p = 2.708×10^−06^) is represented by the horizontal red line, suggestive gene-wide significance (p = 1×10^−03^) is indicated by the blue horizontal line.

### 3.2. Gene and gene-set analyses

Gene-based tests were conducted to test whether any protein-coding gene carries a load of common variation associated with OCS. Using MAGMA v1.08 within FUMA v1.3.7, input SNPs were mapped to 18,464 protein coding genes. No gene reached the Bonferroni-corrected significance threshold (p = 0.05/18,464 = 2.71×10^−06^) in the gene-based test (see Figure 1 for a Miami-plot including the Manhattan-plot of the gene-based test in the lower panel, and Supplementary Figure S14 for a QQ-plot). Further, 15 496 gene sets (Curated gene sets: 5 500, GO terms: 9 996) from MsigDB v7.0 were tested for association. One curated gene set (Mootha Gluconeogenesis) reached Bonferroni-corrected significance (N_genes_ = 28, Beta= 0.7872, SE = 0.1611, P_bon_ = 0.008).

### 3.3. Heritability and cross-disorder analyses

#### 3.3.1. Heritability estimates

For the twin cohorts, the additive genetic variance of OCS, estimated based on the sparse genetic relatedness matrix, ranged between 0.33 (NTR) and 0.43 (TwinsUK), with estimates for the STR cohorts in between: STR-CATSS: 0.35 (GSA chip sub-sample), STR-CATSS: 0.41 (PsychChip sub-sample), STR-YATSS: 0.39, STR-STAGE: 0.39. Note that these heritability estimates based on the twin cohorts are largely driven by the twin resemblance (∼0.5 between DZ twins and siblings, 1.0 for MZ twins, and 0 between unrelated individuals). The heritability for SfS, only including unrelated individuals, was 0.083 (SE = 0.053, P = 0.0516). The SNP-based heritability estimate of the GWAS meta-analysis using LDSC resulted in a total observed scale h² of 0.041 (SE = 0.0144, Z = 2.85, P=0.0044).

#### 3.3.2. Cross-disorder genetic correlations

Of the genetic correlations between OCD and 97 other phenotypes, including psychiatric, personality, psychological, substance-use, neurological, cognition, socioeconomic status, autoimmune, cardio-vascular, anthropomorphic, and fertility traits, 24 exceeded the FDR-corrected significance threshold (Figure 2). As expected, OCS were most strongly associated with OCD out of all psychiatric disorders, followed by anxiety, depressive disorder (DEP), major depressive disorder (MDD), schizophrenia (SCZ), and anorexia nervosa (AN). Significant positive genetic correlations were also observed for all 15 neuroticism phenotypes (14 sub-items and the neuroticism total score). Higher correlations emerged for all worry-related items, and slightly lower correlations for all depressive-related items. A significant positive correlation was further observed for “cigarettes per day” and “tiredness”, while “subjective well-being” yielded a significant negative correlation (see Figure 2, and Supplementary Table S4). We further compared the genetic correlation patterns of OCS and OCD with all other 97 traits, to identify concordant and discordant patterns. There was a clear relationship between the two correlation-patterns of OCS and OCD (see Figure 3), with 66 traits showing concordant directions of correlation (i.e., both correlations above 0 or both below 0), and 30 traits showing a discordant direction of correlation (i.e., one correlation above 0 and one below 0). Especially strong concordance was observed for the psychiatric and personality traits. Of the significant correlations with OCS, only ‘*neuroticism loneliness’* (non-significant) and *‘cigarettes per day’* (significant) showed a different direction of effect with OCD (both positive with OCS, but negative with OCD).

**Figure 2:**
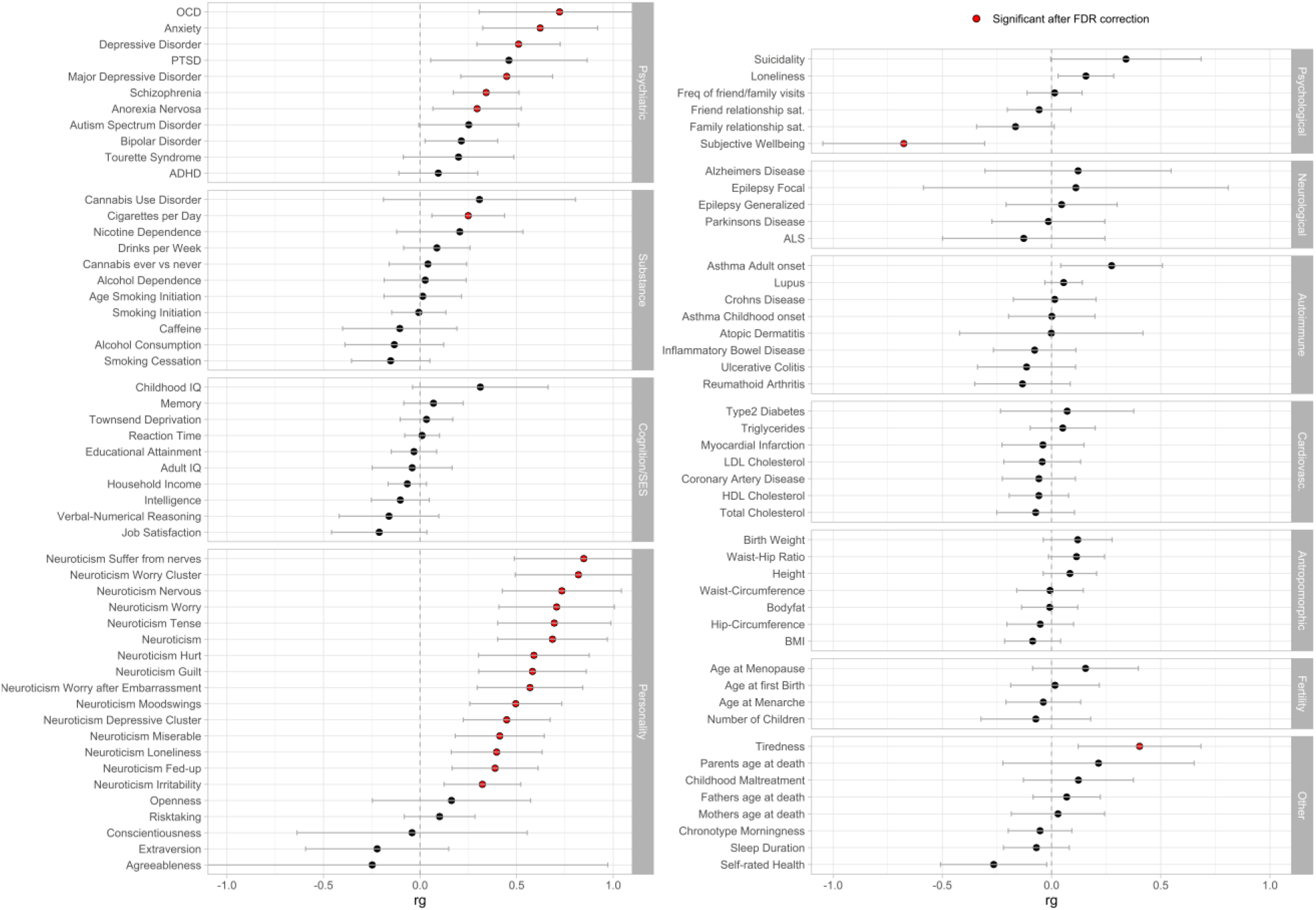
Genetic correlations. (rg) between OCS and a broad range (N = 97) of other phenotypes, assembled into 11 groups (psychiatric, substance, cognition/socioeconomic status (SES), personality, psychological, neurological, autoimmune, cardiovascular, anthropomorphic, fertility, and other). Error bars represent 95% confidence intervals; red circles indicate significant association after FDR correction for multiple testing.

**Figure 3:**
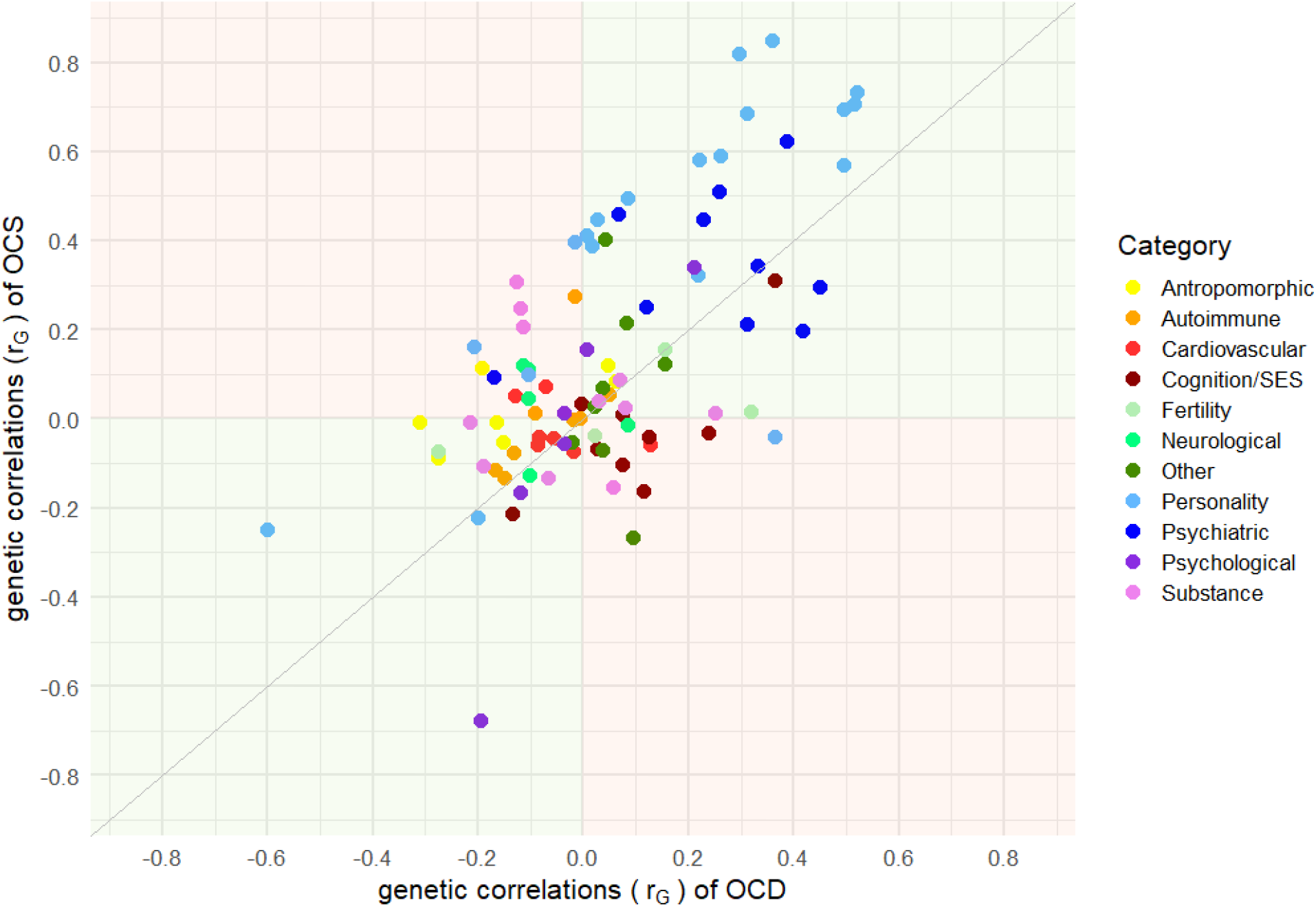
Comparison between genetic correlation estimates (r_G_) of OCS and OCD for 97 other phenotypes, color-coded according to 11 groups (psychiatric, substance, cognition/socioeconomic status (SES), personality, psychological, neurological, autoimmune, cardiovascular, anthropomorphic, fertility, and other). On the x-axis the genetic correlation estimates for OCD are displayed, on the y-axis for OCS. Green quadrants indicate concordance, red quadrants discordance in the direction of genetic correlations between OCS and OCD.

#### 3.3.3. Polygenic risk score analyses

We calculated a range of PRSs, based on publicly available summary statistics of OCD, DEP, SCZ, ASD, ADHD, and EA and examined their relationship with OCS in each cohort. GEE (STR, NTR, TwinsUK) and linear regression analyses (SfS) revealed significant (Bonferroni-corrected p < 0.05/7 = 0.0071) associations between OCS and PRS based on OCD, DEP, SCZ, and EA, but not consistently across all target datasets (see Supplementary Table S4). In the meta-analysis, summarizing the PRS results across all target cohorts, the OCD (P_fixed_ = 3.06 ×10^−5^) and SCZ (P_random_ = 3.69 × 10^−6^) PRS showed significant associations with OCS. The DEP PRS also showed a significant association with OCS, however, there was significant heterogeneity across the four cohorts (P_Q_ = 0.0020), and the random effects model failed to reach significance (P_random_ = 0.0801). The other traits’ PRSs (ASD, ADHD, EA) were not significantly associated with OCS in the PRS meta-analysis (All meta-analyzed results are shown in Table 2).

**Table 2:** The table shows the PRS results for OCS (Leave-one-out), obsessive-compulsive disorder (OCD), depressive disorder (DEP), schizophrenia (SCZ), autism-spectrum disorder (ASD), attention-deficit hyperactivity disorder (ADHD), and educational attainment (EA), meta-analyzed across all target datasets for pre-selected p-value thresholds (P_threshold_). As measures of heterogeneity of PRS associations across all target datasets, Higgin’s I² statistic and the p-value for Cochran’s Q test (P_Q_) are reported. P_fixed_ and P_random_ list the p-values of a fixed and a random-effects model, respectively. A random effects model was only calculated if there was substantial (I² > 0.5 and/or P_Q_ < 0.05) heterogeneity across the datasets. The effective sample size (N_effective_) is summed over the separate target datasets. For STR, NTR, and TwinsUK the effective N was determined based on the actual N (including family members) weighted by the ratio of the squared SE from the GEE sandwich-corrected model and the naive model. For SfS the sample N is listed. Bonferroni-corrected significance threshold was set to 0.00714 (i.e. 0.05/7), significant p-values are in bold.

| Discovery | P<sub>threshold</sub> | N<sub>effective</sub> | Heterogeneity |  | P<sub>fixed</sub> | P<sub>random</sub> |
| --- | --- | --- | --- | --- | --- | --- |
|  |  |  | I<sup>2</sup> | P<sub>Q</sub> |  |  |
| OCS (Leave-one-out) |  |  |  |  |  |  |
| OCS | 0.5 | 30342.69 | 0 | 0.78822 | 5.03 x 10<sup>-7</sup> |  |
| Cross-trait |  |  |  |  |  |  |
| OCD | 0.1 | 31125.74 | 0.1540 | 0.31500 | 3.06 x10<sup>-5</sup> |  |
| DEP | 0.05 | 30245.80 | 0.7972 | 0.00201 | 2.12 x 10<sup>-9</sup> | 0.080143 |
| SCZ | 0.1 | 30817.02 | 0.5667 | 0.07436 | 4.22 x 10<sup>-17</sup> | 3.69 x 10<sup>-6</sup> |
| ASD | 0.1 | 32446.66 | 0.4640 | 0.13300 | 0.04289 |  |
| ADHD | 0.1 | 29644.22 | 0.7391 | 0.011118 | 0.11762 | 0.518405 |
| EA | 1 | 30866.38 | 0.8944 | 2.98 x 10-6 | 0.52699 | 0.793110 |

### 3.4. Compatibility between cohorts

No genome-wide significant heterogeneity was observed in the OCS GWAS meta-analysis (see Supplementary Figure S16 for Manhattan plot and QQplot). The LOO PRS meta-analysis showed a significant positive association between the LOO PRS and OCS (P_fixed_ = 5.03×10^−07^) (see Table 2 for the PRS meta-analysis results and Supplementary Table S4 for individual results for each target cohort).

Given the low power for the main GWAS analysis, the power for LOO GWAS analyses and sign-tests between partial analyses is even lower, making it difficult to draw any definitive conclusions from the results. While no individual study markedly stands out from the rest, results of the sign tests analyses fluctuate with estimates ranging from 0.28 to 0.68 (and between 0 and 1 for P = 1×10^−06^). See Supplementary Figure S17-S20 for Manhattan-plots and QQ-plots of the LOO GWASs and Supplementary Table S5 and S6 for Sign-test results.

We further calculated genetic correlations between each LOO OCS meta-analysis and the same 97 phenotypes (described above) to compare the individual influence of each cohort on the overall correlation estimates. When not considering the individual sub-items of neuroticism, the LOO GWAS excluding SfS showed 13 significant correlations, the GWAS without TwinsUK showed eight significant correlations, the GWAS without STR showed two significant correlations, while the GWAS excluding NTR did not significantly correlate with any of the traits (see Supplementary Figure S21). As sample sizes for the LOO GWAS meta-analyses varied (LOO STR: N = 17 055; LOO NTR: N = 25 393; LOO SfS: N = 28 772; LOO TwinsUK: N = 30 609), it was expected that the power to detect significant correlations for each LOO GWAS differs. For almost all genetic correlations, each LOO GWAS meta-analysis showed the same direction of effect. For all correlations with psychiatric disorders and neuroticism phenotypes, the LOO analysis excluding NTR showed slightly higher estimates (but also larger confidence intervals).

## 4. Discussion

This is the first meta-analysis aiming to identify the genetic underpinnings of OCS in the general population. Though we could not replicate previous findings, two SNPs previously associated with OCS reached suggestive significance in this meta-analysis: the SNP found by den Braber et al (rs8100480, *P* = 2.56×10^−08^ (20)) had a p-value of 0.015 in the current study, while the SNP reported in Burton et al. (rs7856850, *P* = 1.48×10^−08^ (13)) had a p-value of 0. The SNP with the lowest p-value (P = 6.36×10^−08^) in the meta-analysis was rs113538937 on chromosome 4, tagging eight genes which have previously been associated with alcohol use (*HTT*), smoking (*GRK4, NOP14, NOP14-AS1, ADD1, SH3BP2*), worry (*HTT*), and measures of socioeconomic status and education (*HTT, GRK4, NOP14, NOP14-AS, ADD1*). In the gene-based tests, no gene achieved genome-wide significance. However, one gene-set reached Bonferroni-corrected significance. Expression of genes in this set is high at sites of insulin-mediated glucose disposal (61). Dysregulated insulin signaling in the etiology of OCS has been suggested by a previous study describing a genetic overlap of OCS with an insulin signaling-related trait in children and adolescents (62). Epidemiological studies similarly support the role of dysregulated insulin signaling in OCD and OCS. Specifically, patients diagnosed with OCD were found to have a significantly higher risk of developing type 2 diabetes compared to population controls (63).

Our results show that OCS share genetic risk with OCD. Polygenic risk for OCD was associated with OCS and OCS and OCD case/control status showed a substantial genetic correlation (r_G_ = 0.72, p = 0.0007). This is comparable to estimates reported in recent studies (r_G_ = 0.61, p = 0.017 (12); r_G_ = 0.83, p = 0.07 (13)). Notably, OCS did show the highest correlation coefficient with OCD (r_G_=0.72), but the strength of this genetic correlation was statistically not significantly different from that with anxiety (r_G_=0.62), DEP (r_G_=0.51), SCZ (r_G_=0.34), or AN (r_G_=0.30). While we did not exclude participants with any of these diagnoses, the most likely explanation is that these common comorbidities of OCD share genetic underpinnings with OCS. We also observed a high concordance in direction and strength of the correlation patterns of OCS and OCD with other phenotypes. However, given the present sample size, these values must also be interpreted with caution. A fact that is further illustrated by our PRS analyses: scores calculated based on the most recent SCZ GWAS more accurately predicted OCS than scores calculated based on the most recent OCD GWAS, which is grossly underpowered. This suggests that larger cohorts are needed for the accurate estimation of these associations.

SNP-based heritability for OCS in the current sample of 4.1% was significant, but somewhat lower than previous studies (7-16%; 12,13,20). This could possibly be explained by the heterogeneity across the 8 cohorts. Specifically, these samples differed in their ascertainment (twins vs community-based samples), instruments employed, as well as age range of participants. While no specific pattern emerged in our compatibility analyses, the sign tests indicated some level of heterogeneity. In the PRS analyses, prediction results deviated mostly for the SfS sample, which is the youngest and only non-twin sample included. It also uses the only questionnaire that is coded from strengths to weaknesses (from -3 meaning far less often than average to +3 meaning far more often than average), which could have led to further differences between the cohorts. With an estimated 4.1% the SNP-based heritability also stands in stark contrast with heritability estimates observed in twin studies (37-41%; (64)). However, in line with previous research reports, we found a lower SNP-based heritability for OCS (12,13) than for clinical OCD (0.28–0.37) (65,66). The reason for the disparity in SNP heritability between traits and diagnosis is unclear but could be explained by the fact that clinical diagnosed OCD represents the extreme of the OCS distribution in the general population. Previous studies of related psychiatric disorders, such as schizophrenia, revealed that the genetic risk is higher for more severe and chronic cases (67,68). In addition to this consideration of impairment, differences including informant (parent/self vs. clinician), type of measurement (categorical vs. quantitative), and timing (cross-sectional vs. lifetime symptoms) could contribute to the divergence in SNP-based heritability estimates of OCS and OCD.

The present study had several limitations. First, despite being the largest meta-analysis of OCS including, to our knowledge, all samples currently available worldwide, the sample size is still relatively small for estimating heritability and detecting specific significant genetic markers. The meta-analysis also currently lacks the integration of non-European samples. The present results thus call for replication in, and extension to larger and more diverse cohorts. Second, previous research has suggested that OCS dimensions (e.g. contamination, checking, harm or symmetry) may be etiologically heterogeneous (12,62,64,69). As such, future studies might aim to identify the genetic underpinnings of specific OCS dimensions. Third, no information was available on the presence of common clinical comorbidities in all samples. This precluded detailed analysis of the identified genetic overlap. Future studies in larger cohorts should also investigate in more detail how OCS relate to other phenotypes, for instance addiction and personality disorders. Fourth, the observation that patients often establish non-random relationships with persons affected by the same or another mental disorder (70), might extend to people with OCS and contribute to the observed genetic correlations of OCS with anxiety, DEP, SCZ and AN. However, the LD-score method does not investigate the impact of assortative mating (71). Therefore, assessing the degree to which this phenomenon may have influenced the genetic correlation estimates was beyond the scope of the present study. Future investigations of larger data sets for OCS and other psychiatric disorders are needed to refine the analysis of shared and specific genetic risk as well as communalities and specificities of the respective disorders.

To summarize, OCS have a significant polygenic contribution and share genetic risk with diagnosed OCD, supporting the hypothesis that OCD represents the extreme end of widely distributed OCS in the population.

## Data Availability

The meta-analyzed summary statistics will be made available via the Psychiatric Genomics
Consortium Download page (https://www.med.unc.edu/pgc/download-results/).

## 5. Acknowledgements

We acknowledge The Swedish Twin Registry (STR) for data access. The STR is managed by Karolinska Institutet and receives funding through the Swedish Research Council under the grant no 2017-00641. The computations/data handling were enabled by resources provided by the Swedish National Infrastructure for Computing (SNIC) at Uppmax partially funded by the Swedish Research Council through grant agreement no. 2018-05973.

The Netherlands Twin Register (NTR) warmly thanks all twin families for their participation. NTR is supported by multiple grants from the Netherlands Organizations for Scientific Research (NWO) and Medical Research (ZonMW): Netherlands Twin Registry Repository (NWO 480-15-001/674); the Biobank-based integrative omics study (BIOS) funded by BBMRI-NL (NWO projects 184.021.007 and 184.033.111); the European Science Council (ERC) Genetics of Mental Illness (ERC Advanced, 230374, PI Boomsma); the Royal Netherlands Academy of Science Professor Award (PAH/6635) to D.I.B.; Rutgers University Cell and DNA Repository (NIMH U24 MH068457-06), the Avera Institute, Sioux Falls, South Dakota (USA) and the National Institutes of Health (NIH R01 HD042157-01A1). Part of the genotyping was funded by the Genetic Association Information Network (GAIN) of the Foundation for the National Institutes of Health and Grand Opportunity grants 1RC2 MH089951).

TwinsUK is funded by the Wellcome Trust, Medical Research Council, Versus Arthritis, European Union Horizon 2020, Chronic Disease Research Foundation (CDRF), Zoe Ltd, the National Institute for Health and Care Research (NIHR) Clinical Research Network (CRN) and Biomedical Research Centre based at Guy’s and St Thomas’ NHS Foundation Trust in partnership with King’s College London.

Spit for Science was supported by the Canadian Institutes of Health Research (R.J.S., MOP-93696 and P.D.A., MOP-106573). Dr. Arnold is supported by the Alberta Innovates Translational Health Chair in Child and Youth Mental Health.

## 7. Data availability

The meta-analyzed summary statistics will be made available via the Psychiatric Genomics Consortium Download page (https://www.med.unc.edu/pgc/download-results/).

## 8. Author contributions

PDA, DJAS, SMM, DMC, DIB, MM, and NIS contributed to the conception of the over-all study design; CLB, LA, RP, ML, JJC, JJH, VZI, HL, PL, PM, CR, RJS, HMW, JC, PDA, DIB, SMM, DMC, and DC contributed to the data collection of the individual datasets and/or provided code; NIS, DJAS, TS, and CI conducted all primary data analyses; NIS, DJAS, SMM, and CLB drafted the manuscript; all authors provided critical edits and discussions and approved the submitted version of the manuscript.

