## Supplementary Information for "Genome-Wide Association Study of Obsessive-Compulsive Symptoms including 33 943 individuals from the general population"

### Supplementary Material Obsessive-Compulsive Symptom GWAS

**1. Methods** 1
1.1. Cohort descriptions 1
1.2. Distribution of OCS items and sum-scores 3
1.3. Distribution of standardized OCS scores (Z-scores) 12
1.4. Genotyping, quality control and imputation of individual cohorts 14
**2. Results** 18
2.1. Genome-wide association results 18
2.2. Compatibility between cohorts 20

### 1. Methods

#### 1.1. Cohort descriptions

*STR*

Individuals included in this study were monozygotic (MZ) or dizygotic (DZ) twins enrolled in the population-based *Swedish Twin Registry* (STR), who participated in one of its large-scale cohort studies, namely the *Child and Adolescent Twin Study in Sweden* (CATSS18, CATSS24), the *Study of Twin Adults: Genes and Environment* (STAGE), or the *Young Adult Twins in Sweden Study* (YATSS). CATSS is a prospective, longitudinal study of all twins born in Sweden since 1992, in the present study data measured at age 18 (Catss18), and/or age 24 (Catss24) was of relevance (see Anckarsäter et al. (1) for details). CATSS data collection was initiated in 2004. STAGE includes twins born in Sweden between 1958 and 1985, data collection took place between 2005 and 2006. The sample was restricted to twins whose co-twin survived until at least 1 year of age. The YATSS survey took place in 2013, including twin-pairs born between May 1986 and June 1992. For all studies, collected data types include phenotypic and exposure data gathered via questionnaires and interviews (in person, online, and telephone-based), as well as genotypic data through saliva samples. DNA extraction from saliva started in 2009 when kits to extract saliva were sent to the participants after the completion of the online-based or telephone interviews. DNA extraction was then performed at the Karolinska Institutet biobank. See Zagai et al. (2) for details of the STR. All participants gave their informed consent. The study was approved under the reference number 2018/2232-32.

*NTR*

Twins and their family members (parents, children, siblings) registered at the Netherlands Twin Register (NTR) participated. Every two to three years, subjects who are registered receive self-report surveys that contain a variety of questionnaires related to health, personality, demographics, lifestyle and psychiatric disorders (3). Data on OC symptoms were available for 20,438 subjects in the questionnaires sent in 2005 and 2008. Of these, N = 8,550 (64.2% female) had genotype data available and were of European ancestry based on a Mahalanobis distance across the first 5 PCs. The mean age at time of filling out the questionnaire for the last available observation was 41.9 years (SD 15.8; age range 13–82 years). Informed consent was obtained from all participants. The NTR data collection was approved by the Central Ethics Committee on Research Involving Human Subjects of the VU University Medical Centre, Amsterdam, an Institutional Review Board certified by the U.S. Office of Human Research Protections (IRB number IRB00002991 under Federal-wide Assurance - FWA00017598; IRB/institute codes, NTR 03-180).

*SfS*

Spit for Science (SfS) is a population-based cohort consisting of children and adolescents recruited at the Ontario Science Center (OSC) in Toronto, Canada. For descriptions of the SfS cohort, refer to Crosbie et al. (4). Here, we analyzed 5,171 unrelated SfS participants of self-report European ancestry (confirmed using genetic data) who provided complete demographic and OCS questionnaire information. This information was collected over a 16-month period between 2008 and 2009. Protocols for obtaining informed consent and assent (where applicable) were approved by the Research Ethics Board at the Hospital for Sick Children. All participants provided saliva samples for genotyping.

*TwinsUK*

Participants were monozygotic and dizygotic twins from the TwinsUK adult twin registry (www.twinsuk.ac.uk). The characteristics of the sample are described elsewhere (5). This registry data set consists of approximately 10,000 monozygotic and dizygotic twin pairs of European ancestry, from the United Kingdom, aged 16 and above. Recruitment was through a series of media advertisements that did not target individuals on the basis of a pre-existing disorder. The characteristics of the cohort have been shown to be comparable with age-matched population singletons in terms of disease-associated and lifestyle characteristics (6). Zygosity status was initially obtained using the "Peas in the Pod" questionnaire (7) and has been further validated using genome-wide genetic data. The Obsessive Compulsive Inventory (8) was sent to all active twins in the registry (N = 8,313) as part of a larger wave of data collection. Analyzable questionnaire data was returned by 5,154 twins. Hence the full-size UKTwins Obsessive-Compulsive data set consists of 4,932 twin pairs (161 monozygotic males, 162 dizygotic males, 1,172 monozygotic females, 953 dizygotic females, and 108 dizygotic twins of the opposite sex, 2 sets of MZ triplets) and 296 singleton twins.

#### 1.2. Distribution of OCS items and sum-scores

STR

**Supplementary Figure S1:** Distribution of all 12 raw OC items of the Brief Obsessive-Compulsive Scale (BOCS) (A-L) in STR CATSS18-GSA. 0 indicates that the symptom was never present, 1 indicates that the symptom was (past) or is present (current). Sum-score (M) consists of the summation of all 12 items.

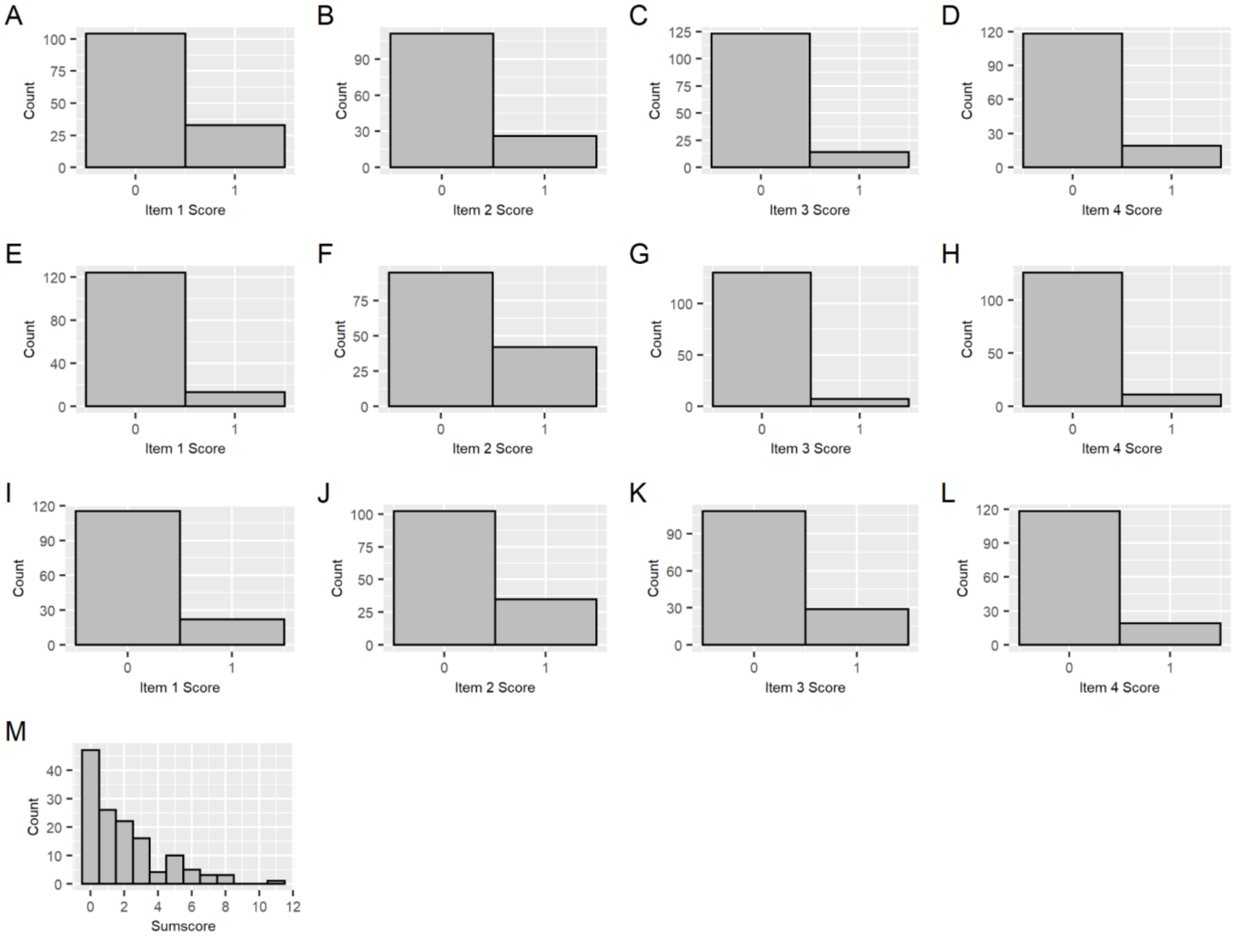

**Supplementary Figure S2:** Distribution of all 12 raw OC items of the Brief Obsessive-Compulsive Scale (BOCS) (A-L) in STR CATSS18-PC. 0 indicates that the symptom was never present, 1 indicates that the symptom was (past) or is present (current). Sum-score (M) consists of the summation of all 12 items.

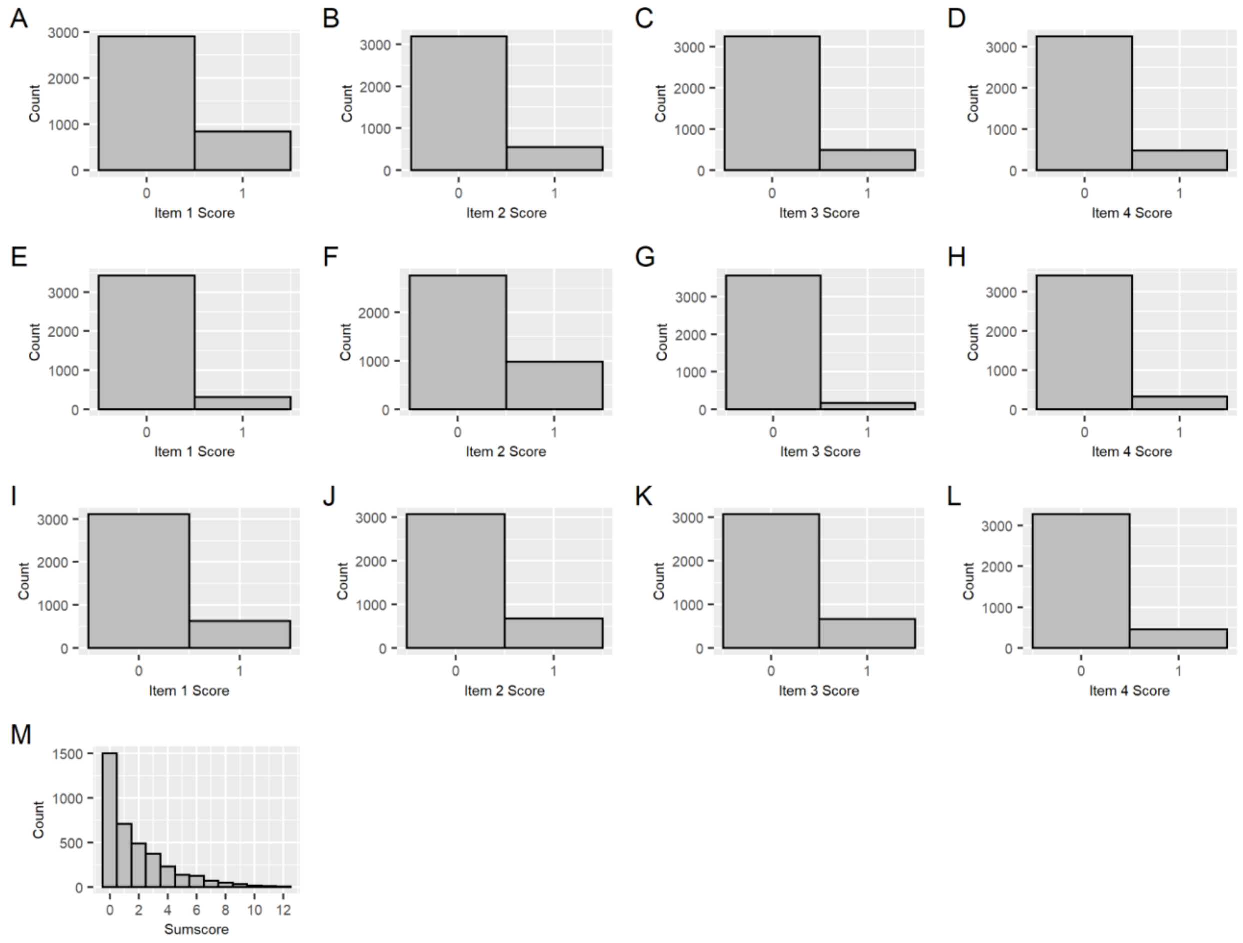

**Supplementary Figure S3:** Distribution of all 12 raw OC items (A-L) of the Obsessive-Compulsive Inventory Revised (OCI-R), excluding hoarding and neutralizing scales in STR CATSS24-GSA. A score of 1 indicates “not at all”, of 2 “a little”, of 3 “moderately”, of 4 “a lot”, and of 5 “extremely”. Sum-score (M) consists of the summation of all 12 items.

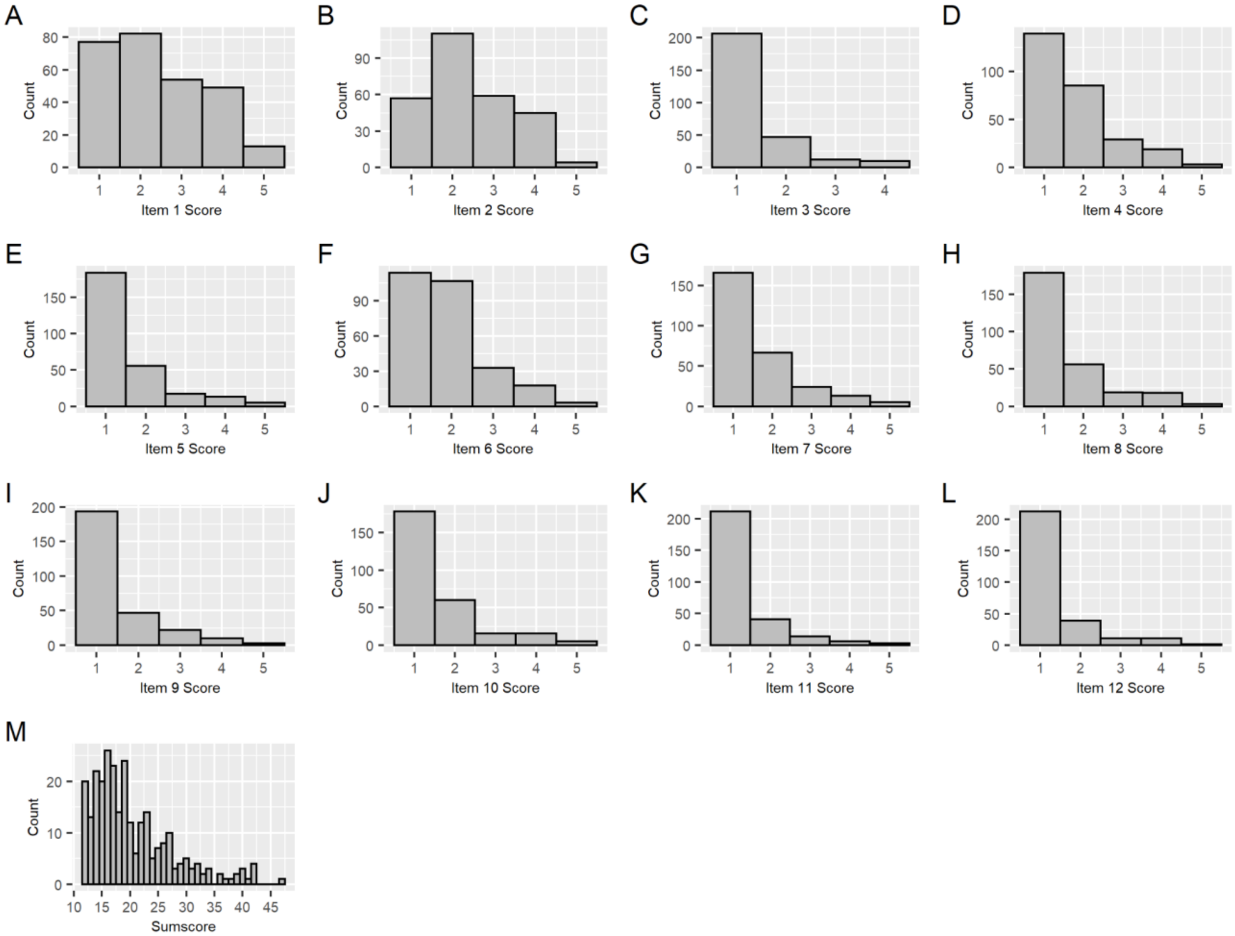

**Supplementary Figure S4:** Distribution of all 12 raw OC items (A-L) of the Obsessive-Compulsive Inventory Revised (OCI-R), excluding hoarding and neutralizing scales in STR CATSS24-PC. A score of 1 indicates “not at all”, of 2 “a little”, of 3 “moderately”, of 4 “a lot”, and of 5 “extremely”. Sum-score (M) consists of the summation of all 12 items.

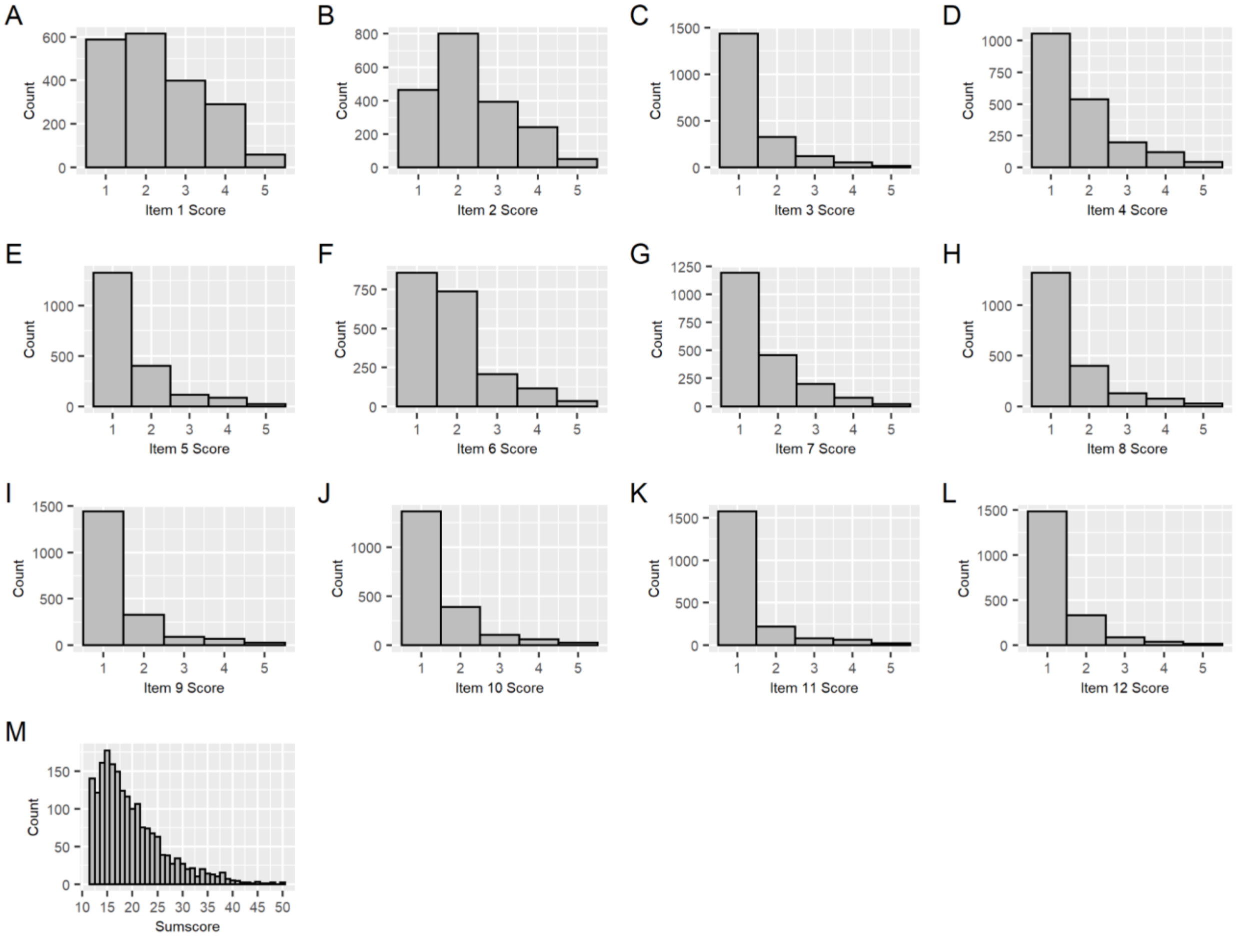

**Supplementary Figure S5:** Distribution of all 7 raw OC items (A-G) in STR STAGE. A score of 1 indicates “symptom not present”, of 2 “a little”, and of 3 “a lot”. Sum-score (H) consists of the summation of all 12 items.

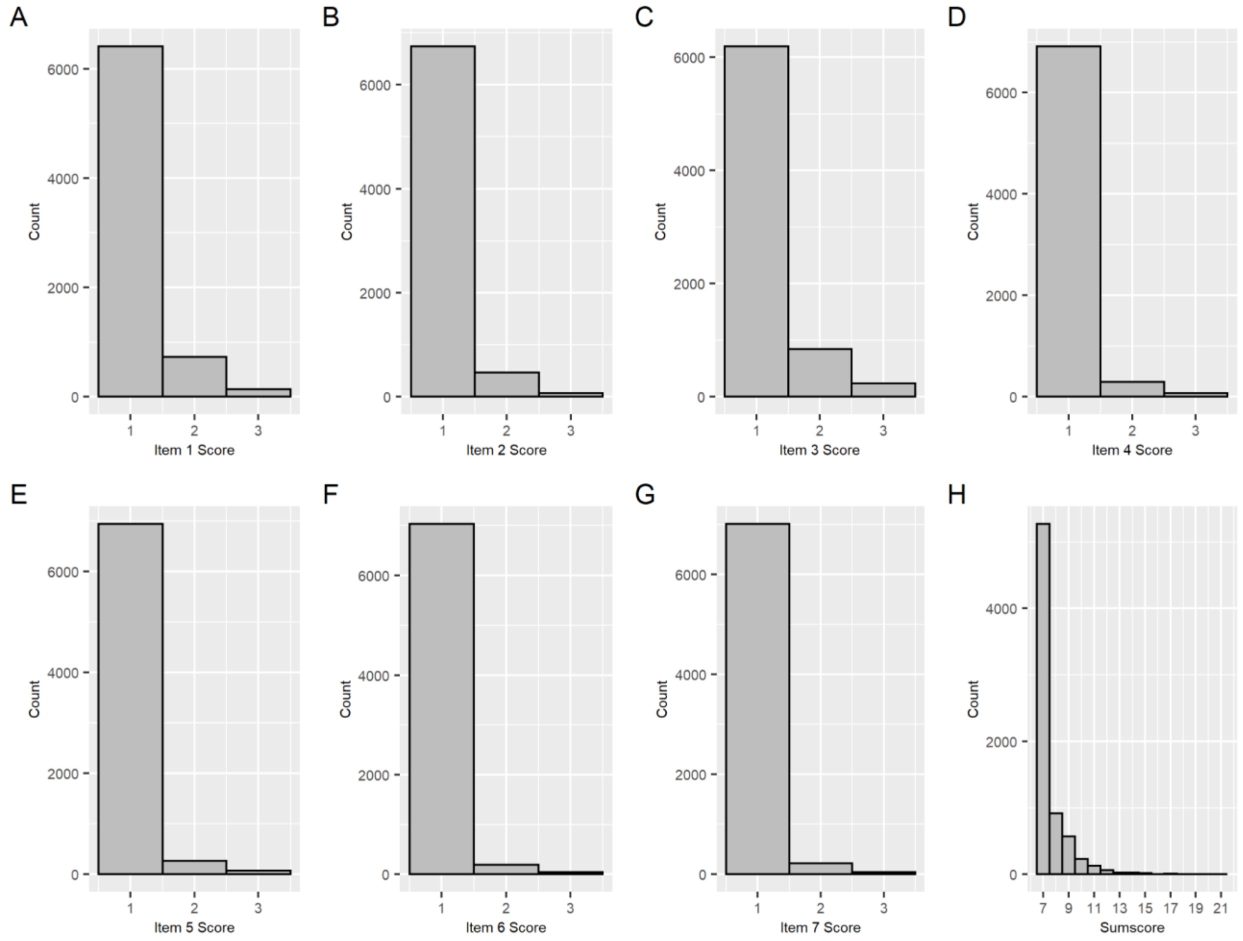

**Supplementary Figure S6:** Distribution of all 12 raw OC items (A-L) of the Obsessive-Compulsive Inventory Revised (OCI-R), excluding hoarding and neutralizing scales in STR YATSS. A score of 1 indicates “not at all”, of 2 “a little”, of 3 “moderately”, of 4 “a lot”, and of 5 “extremely”. Sum-score (M) consists of the summation of all 12 items.

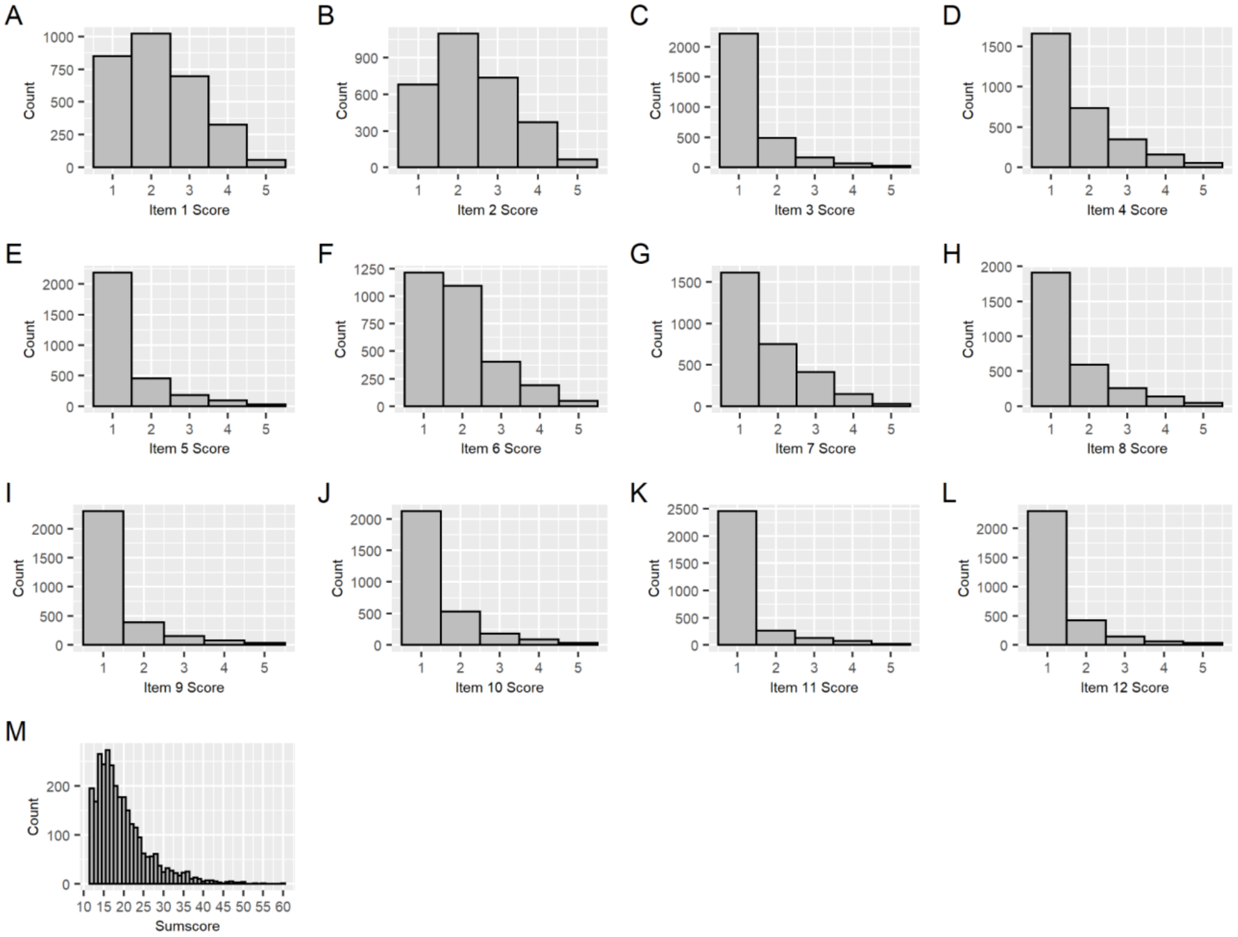

*NTR*

**Supplementary Figure S7:** (**A-I)** Distribution of 9 raw obsessive-compulsive items of the Dutch translation of the Padua Inventory Revised – abbreviated (twelve item original set with rumination items removed). Scoring was on a five-point scale with values 'never', 'very rarely', 'sometimes', 'often','very often'. **(J)** Histogram of the sum-score.

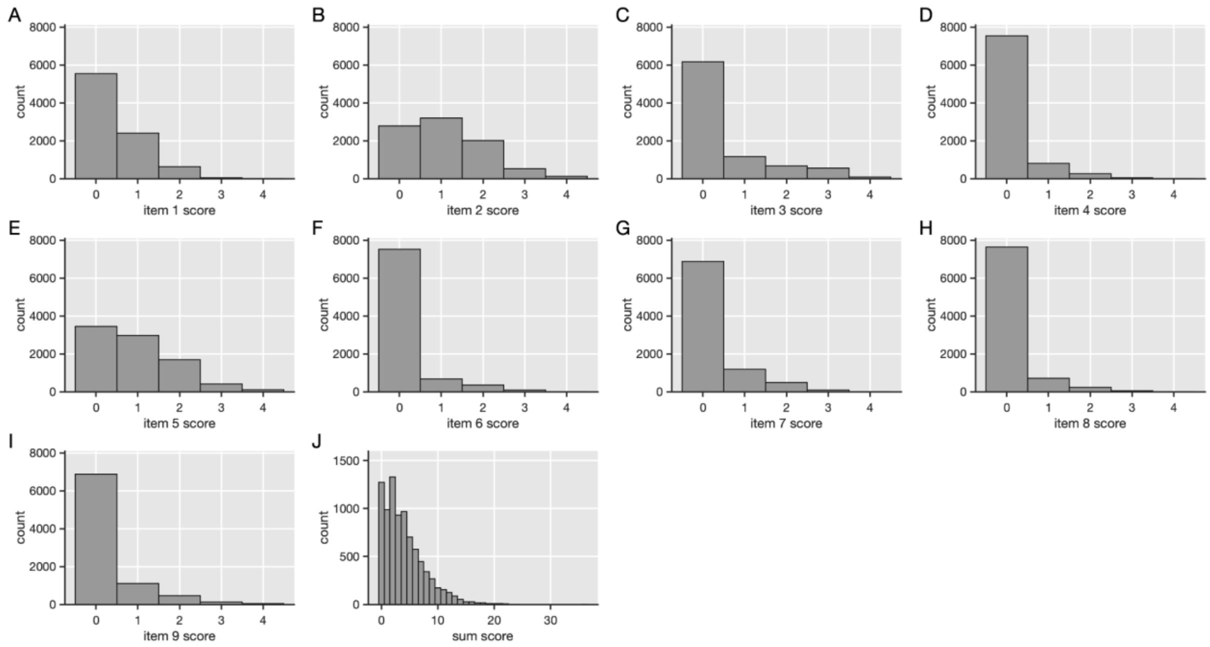

*SfS*

**Supplementary Figure S8:** Distribution of all 19 raw obsessive-compulsive items of the Toronto Obsessive-Compulsive Scale (A-S) in Spit for Science. Total score (T) consists of the summation of all 19 items.

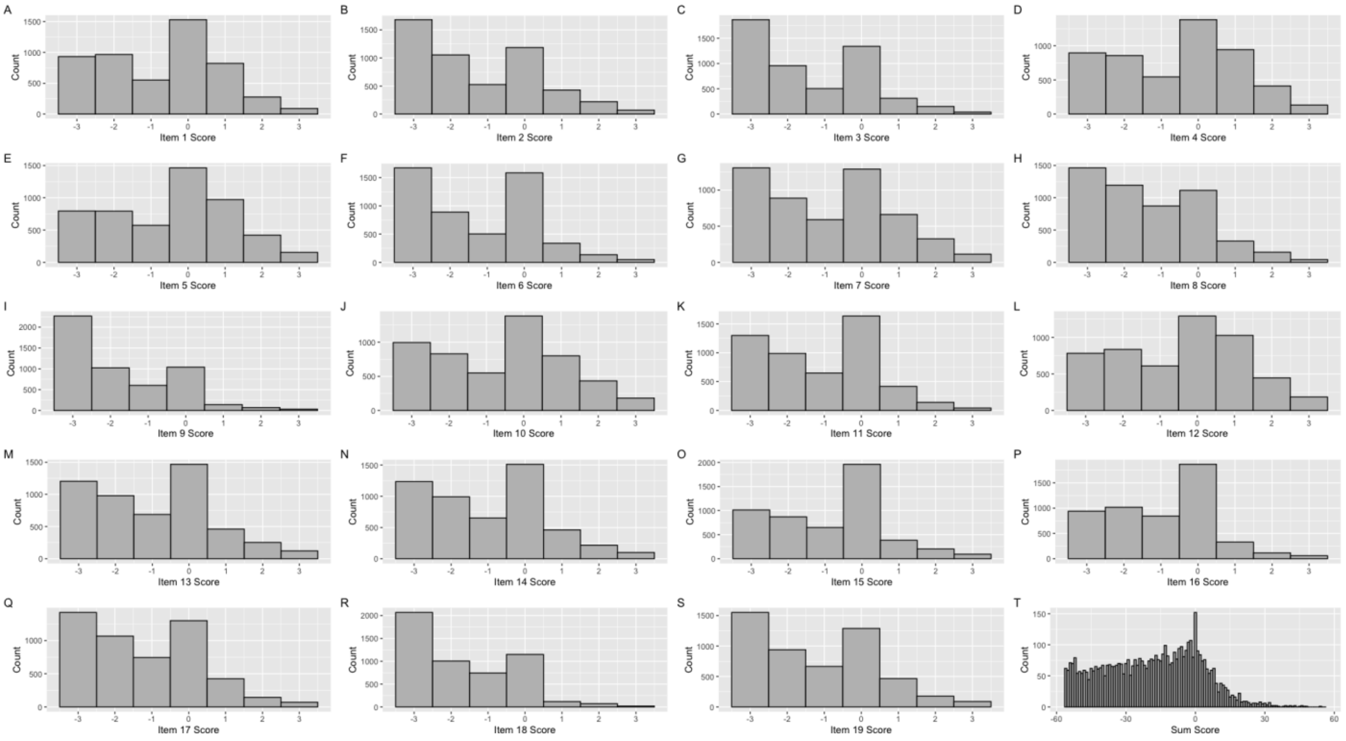

*TwinsUK*

**Supplementary Figure S9:** Distribution of all 12 raw OC items (A-L) of the Obsessive-Compulsive Inventory Revised (OCI-R), excluding hoarding and neutralizing scales in TwinsUK. A score of 1 indicates “not at all”, of 2 “a little”, of 3 “moderately”, of 4 “a lot”, and of 5 “extremely”. Sum-score (M) consists of the summation of all 12 items.

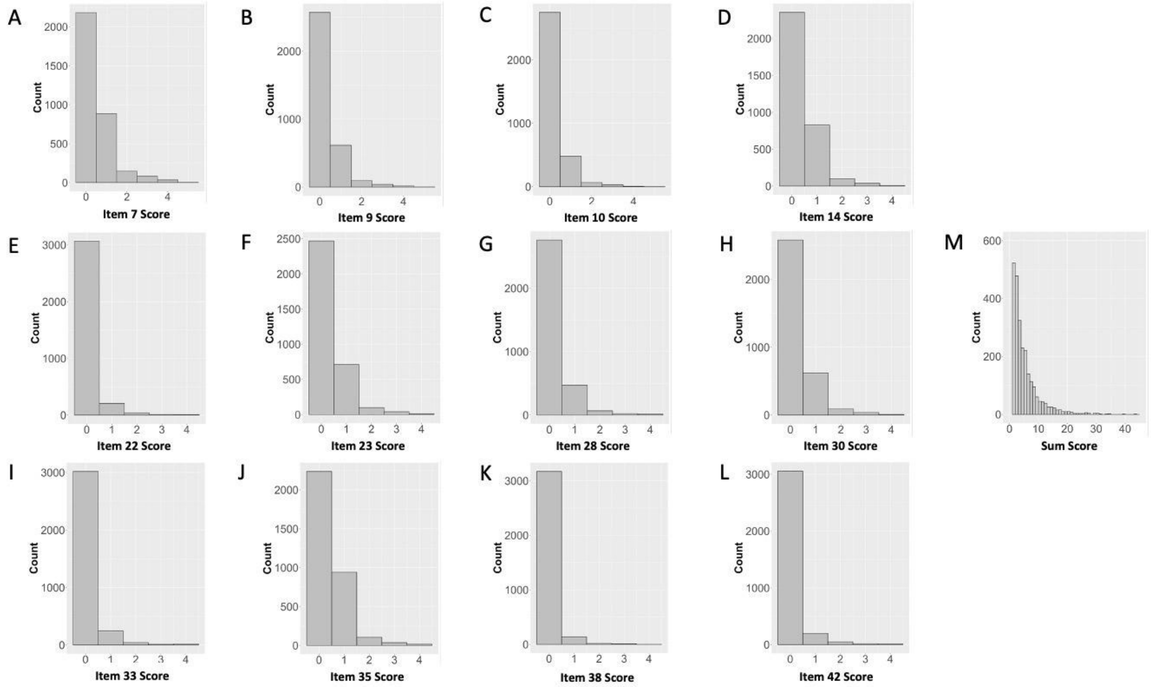

#### 1.3. Distribution of standardized OCS scores (Z-scores)

*STR*

**Supplementary Figure S10:** Distribution of the standardized sum-score (Z-scores) with a mean of 0 and a variance of 1 for each of the STR cohorts that were analyzed in a separate GWAS: Catss1824-GSA (A), Catss1824-PC (B), Stage (C), Yatss (D).

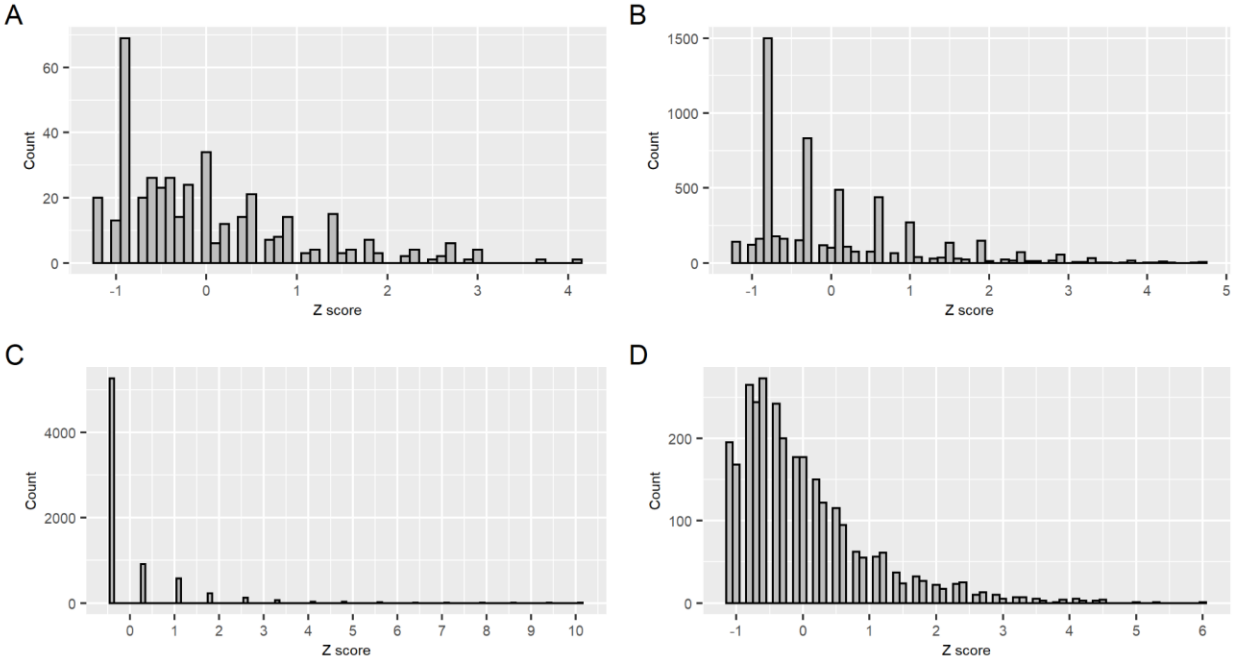


*NTR*

**Supplementary Figure S11:** Distribution of the standardized sum-score (Z-scores) for the NTR sample.

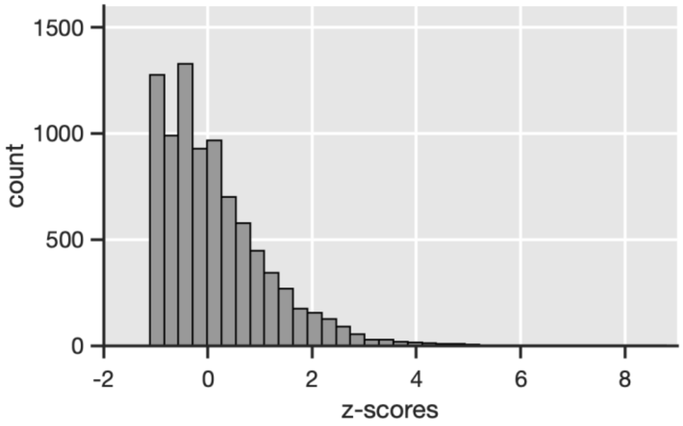

*SfS*

**Supplementary Figure S12:** Distribution of standardized sum-score of the obsessive-compulsive items from the Toronto Obsessive Compulsive Scale in the Spit for Science sample.

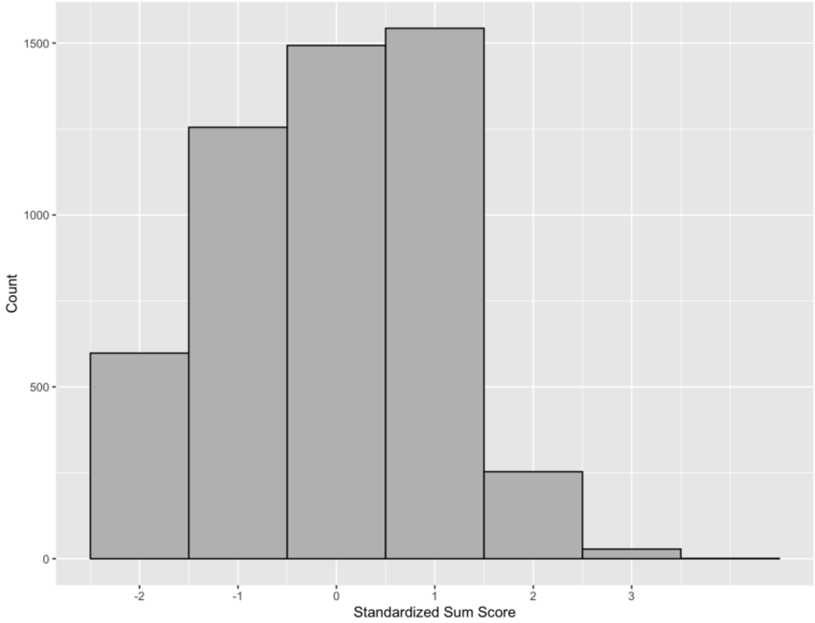


*TwinsUK*

**Supplementary Figure S13:** Distribution of the standardized sum-score (Z-scores) with a mean of 0 and a variance of 1 for the TwinsUK sample.

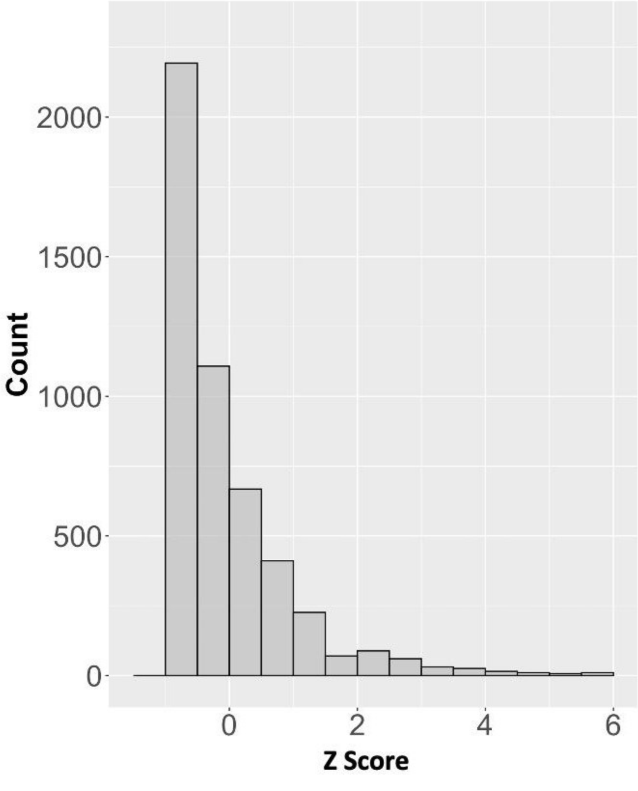

#### 1.4. Genotyping, quality control and imputation of individual cohorts

*STR*

The STR data consists of six different cohorts (Catss18-GSA, Catss24-GSA, Catss18-PC, Catss24-PC, Stage, and Yatss), that were genotyped and imputed in four cohorts (Catss1824-GSA, Catss1824-PC, Stage, and Yatss).

Catss-PC (PsychChip) genotyped samples were processed using the Ricopili pipeline (9) for quality control (QC). In a first round of QC, SNPs with a missingness higher than 0.05 (N = 4,477) were removed, followed by the exclusion of 141 samples due to either per-sample call rate < 0.98, excessive heterozygosity (FHET outside +/- 0.2), or sex mismatch. 146,755 out of 588,454 markers failed SNP QC due to either per-SNP call rate < 0.98, Hardy-Weinberg disequilibrium ($P < 1x10^{-06}$ in MZ twins and $P < 1x10^{-10}$ in DZ twins), or difference in call rate between MZ twins and DZ twins > 0.02. Finally, 139,072 SNPs with minor allele frequency (MAF) < 0.01 were excluded, leaving 302,627 SNPs for principal component (PC) analysis. The first two PCs of the Catss-PC samples were projected onto the 1000 Genomes global population reference panel. 236 samples exceeded six standard deviations from the mean values of the European samples in the 1000 Genomes reference, thereby identifying these samples as non-European ancestral outliers. After the removal of the ancestral outliers, a second round of the QC procedure described above was performed, excluding a further seven samples and 23 SNPs. 10,789 samples and 302,604 SNPs, of which 293,590 were successfully aligned to the forward genomic strand and matched to the reference panel, were then used for imputation. The Sanger imputation server was used to impute the post-QC genotype data, using the Haplotype Reference Consortium (HRC v.1.1) as a reference. EAGLE2 (10) was used for pre-phasing and PBWT was used for imputing. After imputation, 40 million SNPs were available.

Catss-GSA, Stage, and Yatss genotypes were generated each in six batches during 2018 and 2019 on the Illumina Infinium assay (chip GSAMD-24v1-0_20011747_A1) at the SNP&SEQ Technology Platform at Uppsala University using GenomeStudio 2.0.3. 14 (Catss-GSA), 86 (Stage), and six (Yatss) samples were excluded for having sex chromosome abnormalities, genotypic sex different from register information or showing unexpected relatedness patterns, indicating a sample mixup. Data from genotyped monozygotic twin pairs with genotype missingness < 0.02 were merged into one sample per pair (Catss-GSA: N = 160; Stage: N = 54; Yatss: N = 1). 3,898 (Catss-GSA), 8,356 (Stage) and 2,719 (Yatss) genotyped samples were processed using Ricopili. SNPs with a missingness higher than 0.05 (Catss-GSA: N=11,299, Stage: N = 12,831, Yatss: N = 8,076) were removed, followed by the exclusion of 8 (Catss-GSA), 13 (Stage), and 2 (Yatss) samples due to either per-sample call rate < 0.98, excessive heterozygosity (FHET outside +/- 0.2), or sex mismatch. 199,312 out of 700,078 markers (Catss-GSA), 207,658 out of 700,078 makers (Stage), and 199,113 out of 700,078 markers (Yatss) failed SNP QC due to either per-SNP call rate < 0.98, MAF < 0.01, Hardy-Weinberg disequilibrium ($P < 1x10^{-06}$ in MZ twins and $P < 1x10^{-10}$ in DZ twins), or difference in call rate between MZ twins and DZ twins > 0.02. The least common variant was removed for multiallelic sites encoded as multiple markers with the same position but different alleles. Markers having the same position and alleles were merged into one marker per position, removing 625 SNPs for Catss-GSA, 581 for Stage, and 661 SNPs for Yatss, leaving 500,141 (Catss-GSA), 491,839 (Stage), and 500,304 (Yatss) directly genotyped SNPs for analysis. The first two PCs of each study sample were projected onto the 1000 Genomes global population reference panel (11) using plink 1.9 (12). 233 (Catss-GSA), 28 (Stage), and 63 (Yatss) samples exceeded six standard deviations from the mean values of the European samples in the 1000 Genomes reference, thereby identifying these samples as non-European ancestral outliers. Post-QC genotype files were imputed using the Sanger imputation server, using the Haplotype Reference Consortium (HRC v.1.1) (13)as a reference.

EAGLE2 was used for pre-phasing and PBWT was used for imputing. After imputation, 40,359,612 SNPs were available for each dataset. PC analysis without an external population reference was performed to generate ancestry covariates for association analysis. The first 20 PCs based on common (MAF =>0.05) genotyped markers in linkage equilibrium (LD; pairwise $R^2$ <= 0.1), excluding known regions of long-range LD, were derived from unrelated individuals, and then projected on the full sample of twins. Relatedness of individuals within (twin pairs) and between different family IDs was estimated using the KING algorithm in PLINK 2.0 (14). For DZ twins the expected value was 0.25, for MZ twins 0.5. After QC and imputation, MZ co-twins were imputed from their genotyped siblings. The total sample size including MZ co-twins is 4,824 (Catss-GSA), 9,701 (Stage), and 3,358 (Yatss).

We further conducted a combined principal component analysis (PCA) across all four (Catss-GSA, Catss-PC, Stage, Yatss) STR datasets using Ricopili to detect spurious relatedness across the cohorts. One individual of each pair with a pihat > 0.2 was excluded. The number of individuals passing genotype- and phenotype QC was 5,683 (Catss-PC), 412 (Catss-GSA), 7,846 (Stage), and 2,947 (Yatss).

*NTR*

Genotyping was done on multiple platforms over time, namely Perlegen-Affymetrix, Affymetrix 6.0, Affymetrix Axiom, Illumina Human Quad Bead 660, Illumina Omni 1M and Illumina GSA. On each platform genotyping was performed following manufacturers protocols, using the then appropriate calling software. The SNPs of the Perlegen-Affymetrix, Illumina Human Quad Bead 660 and Illumina Omni 1M arrays, which were originally typed on older genome builds, were lifted over to Build 37 HG19 based on RSid locations of the DBSNP 142 marker map. For each genotype platform, samples were removed if DNA sex did not match the expected phenotype, if the Plink heterozygosity F statistic was < -0.10 or > 0.10, or if the genotyping call rate was < 0.90. SNPs were removed if the minor allele frequency (MAF) < 0.01, if the Hardy-Weinberg Equilibrium (HWE) p-value < $1\times10^{-05}$, if call rate < 0.95, or if the N Mendel errors > 20. The absolute value of 20 is used here to remove only the worst offending SNPs (N = 2,902) in platforms that have familial data present (later this is re-filtered more stringently). In addition, palindromic AT/GC SNPs with a MAF range between 0.4 and 0.5 were removed to avoid possible strand alignment issues. For each platform, the data was then position - and strand aligned with the GONL reference set V4. SNPs that had a difference in allele frequency > 0.10 or had mismatching alleles with this reference panel were removed in this step.

The data of the 6 platforms was merged into a single dataset keeping all QCed SNPs of each platform (N=1,781,526). For each individual only one platform was chosen in the following order: Axiom (3,144) > Affy6 (8,640) > 1M (238) > 660 (1,439) > GSA (5,938) > Affy-Perl (1,238). Based on the ~10.6k SNPs that all platforms have in common, DNA IBD was estimated for all individual pairs using the PLINK and KING programs (12,15). These estimates were then compared to the expected familial relations, and samples were removed if these failed to fit. A similar approach was used for DNA zygosity mismatches. Duplicate monozygotic twins N = 3,032, triplets N = 7 as well as NTR samples present in the GONL data N = 364 (plus their MZ-twins) were removed from the data prior to imputation. The data were then cross-platform phased and imputed using MACH-ADMIX to predict the missing SNP genotypes in each platform as compared to the other platforms, based on the complete GONL reference panel haplotypes for the SNPs that were present in at least one platform (the ~1.78m) (16–19). Post imputation, the 2nd (and 3rd) MZ, plus the GONL samples were re-added duplicating the data from the 1st imputed MZ twin, and the complete SNP data from the GONL reference panel.

After this imputation, SNP QC was redone, now using the full merged dataset with all missing genotypes imputed. SNPs were removed if the HWE p-value was < $1\times10^{-5}$, Mendel error rate was more than mean + 3sd, the $R^2$ imputation quality metric was < 0.90 if p-value for association with a single platform vs. all others was < $1\times10^{-5}$. No MAF filter was re-applied (min = 0.0025). This left a cleaned merged dataset of 21,001 NTR individuals with 3,032 MZ pairs, 7 MZ trios, and 1,314,639 SNP markers ($N_{chrX}$ = 20,792). This cross-platform imputed set described is what we consider to be our 'genotyped' dataset in the next two steps, which are the detection of ethnic outliers and the imputation to the 1000 genomes Phase 3v5 and the Human Reference Consortium (HRC) panels (11,13).

Ancestry outliers (non-Dutch ancestry) were defined based on Principal Components Analysis (PCA) by projecting 10 PCs from 1000G reference set populations on the NTR cross-platform imputed data using the SMARTPCA program as described earlier (20,21). Individuals with PC values located outside of the range of European and/or British populations were defined as outliers (N = 1,823). Upon exclusion of outliers, 10 PCs were recomputed for NTR cross-platform imputed data to capture the variation within the Netherlands.

Genotype imputation to the HRC 1.1 (~40m SNPs) and 1000G Phase 3 version 5 (~49m SNPs) reference panels was done on the Michigan Imputation server on the cross-platform imputed data (22). For each reference panel the data were aligned using the PERL based "HRC or 1000G Imputation preparation and checking" tool v4.2.5 (https://www.well.ox.ac.uk/~wrayner/tools/). The remaining SNPs (1,302,481: 1000G, 1,307,940: HRC) were then phased with EAGLE for the autosomes, and SHAPEIT for chromosome X and then imputed using Minimach 3 following the standard imputation procedures of the server (23,24).

The cross-chip imputed data was filtered for SNPs having a MAF < 0.01. Then Genetic Relationship Matrices (GRM) were computed for each individual chromosome (1 to 22) using the GCTA software (25). Subsequently, these 22 matrices were merged into single autosomal matrix. Leave One Chromosome Out (LOCO) GRMs in order to control for genetic background when running genome wide associations, were calculated likewise by merging 22 subsets of 21 chromosomes respectively. Finally, a specific family based GRM was made by setting all individual pairs sharing less than 0.05 of their genomes to 0 in the matrix and leaving the other pairs as calculated (26). No GRMs were calculated for the imputed data (100G and HRC) as these are sufficient to control for confounding in GWAS and to calculate the heritability for various traits.

##### *SfS*

Detailed methods for sample collection, DNA extraction, quantification and genotyping, as well as QC can be found in (27). Briefly, participants' saliva samples were collected using the Oragene OG-500 saliva kits (DNA Genotek, Ottawa, Canada). DNA extraction followed standard methods and was quantified using real-time polymerase chain reaction (rt-PCR). DNA was genotyped on either the Illumina HumanCoreExome or HumanOmni1 beadchip arrays (Illumina, San Diego, CA, USA). Quality control of genetic data was performed in GenomeStudio® and using PLINK (12). Imputation was performed separately depending on batch and Illumina platforms housing Beagle v4.1 (28) and the phase 3 version 5 data of the 1000 Genomes project as a reference. Subsequent analyses were performed using hard called genotypes with imputation quality (INFO) score > 0.8. All participants of non-European ancestry were excluded based on principal components analysis (PCA), and only one participant was included from each family (i.e., the first enrolled sibling). For additional exclusion criteria, refer to (27).

Ancestry related covariates were generated using PCA. PCA was performed without an external population reference using a set of SNPs pruned based on linkage disequilibrium (LD; pairwise $r^2$ < 0.1) and excluding long-range LD regions. To further account for cryptic relatedness in the SfS sample, principal components (PCs) were computed in unrelated individuals and then projected onto the full SfS cohort to yield projected PCs. For subsequent analyses, PCs 1-3 and projected PCs 1-3 were included as covariates.

*TwinsUK*

Genotyping of the TwinsUK dataset was done using the Illumina arrays HumanHap300, HumanHap610Q, 1M-Duo and 1.2MDuo 1M. The normalized intensity data for each of the three arrays was pooled separately, with 1M-Duo and 1.2MDuo 1M pooled together. The Illuminus calling algorithm was used to assign genotypes in the pooled data for each dataset. No call was assigned for genotypes with posterior probabilities below a threshold of 0.95. Validation of pooling was achieved via a visual inspection of 100 random, shared SNPs for overt batch effects. Intensity cluster plots of significant SNPs were visually inspected for over-dispersion, biased no-calling, and/or erroneous genotype assignment. SNPs exhibiting any of these characteristics were discarded.

Quality control steps were implemented using PLINK 1.9. Similar exclusion criteria were applied with respect to (i) HumanHap300 and (ii) HumanHap610Q, 1M-Duo, 1.2MDuo 1M genotyped samples). The following sample-wise exclusion criteria were applied: sample call rate < 98%; heterozygosity across all SNPs ≥ 2 SD from the sample mean; evidence of non-European ancestry (this was assessed by PCA comparison with 1000 Genomes by projecting the first two PCs for UKTwins samples onto the 1000 genomes reference populations, using the EIGENSOFT package); observed pairwise IBD probabilities suggestive of sample identity errors. Misclassified monozygotic and dizygotic twins were corrected based on the derived IBD probabilities, calculated using the KING algorithm in PLINK. Ancestry covariates (20 PCs based on MAF => 0.05 and pairwise $R^2$ <= 0.1 after exclusion of long-range LD regions), were derived for the association analysis for unrelated individuals, by re-calculating PCs after having excluded the PC outliers. Pairwise relatedness of individuals was estimated using the KING algorithm in PLINK 2.0 (15), using expected values of 0.25 for DZ twins and 0.5 for MZ twins. The following SNP-wise exclusion criteria were applied: Hardy-Weinberg p-value < $10^{-06}$, (assessed using unrelated samples); MAF < 1%, in unrelated samples (iii) SNP call rate < 97% (SNPs with MAF ≥ 5%) or < 99% (for 1% ≤ MAF < 5%). Genotypic data resulting from quality control arms (i) and (ii) were combined and the map was converted from Build36 to Build37.
The HRC/1KG Imputation Preparation and Checking Tool (http://www.well.ox.ac.uk/~wrayner/tools/HRC-1000G-check-bim.v4.2.5.zip) (developed by Will Rayner) was used to check input data for accuracy relative to expected 1000G inputs prior to imputation. This process identified errors in the original data, including incorrect REF/ALT designations, incorrect strand designations, extreme deviations from expected allele frequencies, and palindromic (A/T and G/C) SNPs with allele frequency near 0.5 that are often the source of imputation errors. The problematic variants identified were fixed or removed.
The cleaned/updated binary files (one for each chromosome) generated by this tool were used for phasing, using EAGLE for the autosomes and SHAPEIT for chromosome X. They were then imputed using Minimach 3 following standard procedures (23,24). The 1,000 Genomes Phase 3 integrated variant set (NCBI build 37/hg19 coordinates) served as the reference panel. Hard called genotypes with imputation quality (INFO) score > 0.8 were retained for the downstream analyses.

### 2. Results

#### 2.1. Genome-wide association results

**Supplementary Figure S14:** A) The QQ-plot displays quantiles of the -log(10) p-values, resulting from the inverse weighted GWAS meta-analysis, plotted against the quantiles expected under the null hypothesis. 95% confidence interval is indicated by the grey shading. The genomic inflation factor Lambda is the observed median Chi² test statistic under the null hypothesis. Lambda1000 indicates the lambda if the sample contained 1000 individuals. Number of SNPs (N(pvals)) and number of individuals (N(individuals)) included in the meta-analysis are listed. B) The QQ-plot from the gene-based analysis, displaying quantiles of the -log(10) p-values plotted against the quantiles expected under the null hypothesis.

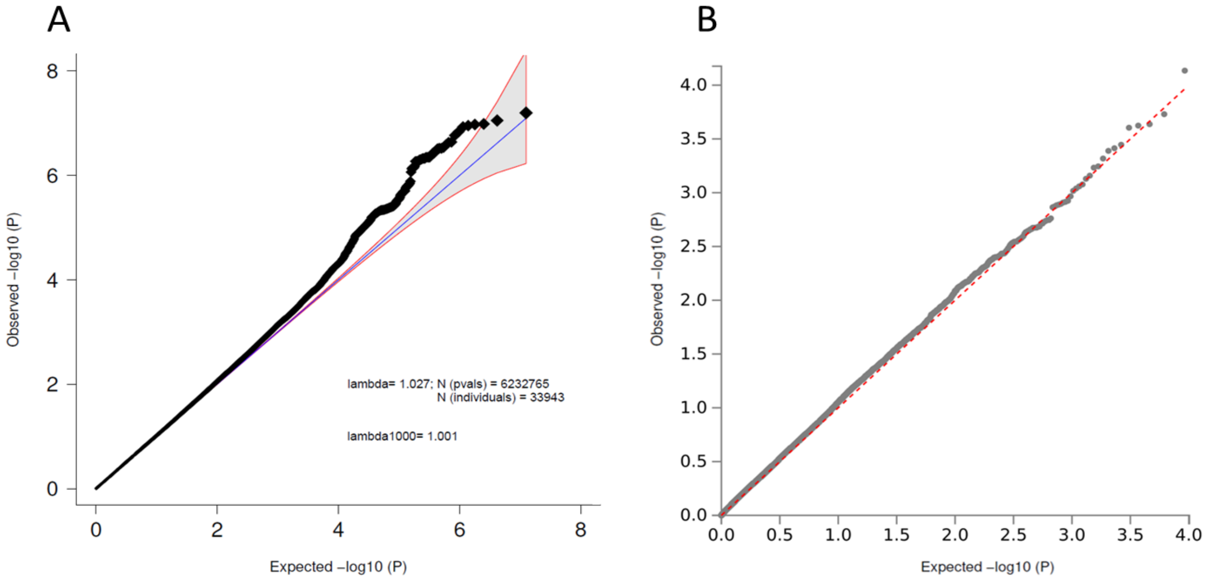

**Supplementary Figure S15:** Forestplot (A) and regional association plot (B) of SNP rs113538937. A) The forestplot shows the effect estimate with 95%-confidence interval for each cohort contributing to the meta-analysis and for the inverse variance weighted meta-analysis (bottom line). The table lists INFO (imputation quality score), p-value (p-value of the SNP effect), f_ca(n) (frequency cases), f_co(n) (frequency controls) (as the current study has a quantitative phenotype, f_ca(n) and f_co(n) are both the same and the overall frequency), ln(OR) (Beta estimate of the effect of the SNP), and STDerr (standard error of ln(OR)) for each of the contributing cohorts and for the meta-analysis. At the top, '+' indicates a positive direction of effect, '-' a negative direction of effect, while '?' indicates that the SNP was not contained in the respective cohort. B) The –log10(p-value) of SNPs in the OCS meta-analysis GWAS is shown on the left y axis. The recombination rates expressed in centimorgans (cM) per Mb (Megabase) (light blue line) are shown on the right y axis. Position in Mb is on the x axis. Only the SNPs with an association p-value less than 0.1 were plotted. The most associated SNP is shown as a purple diamond.

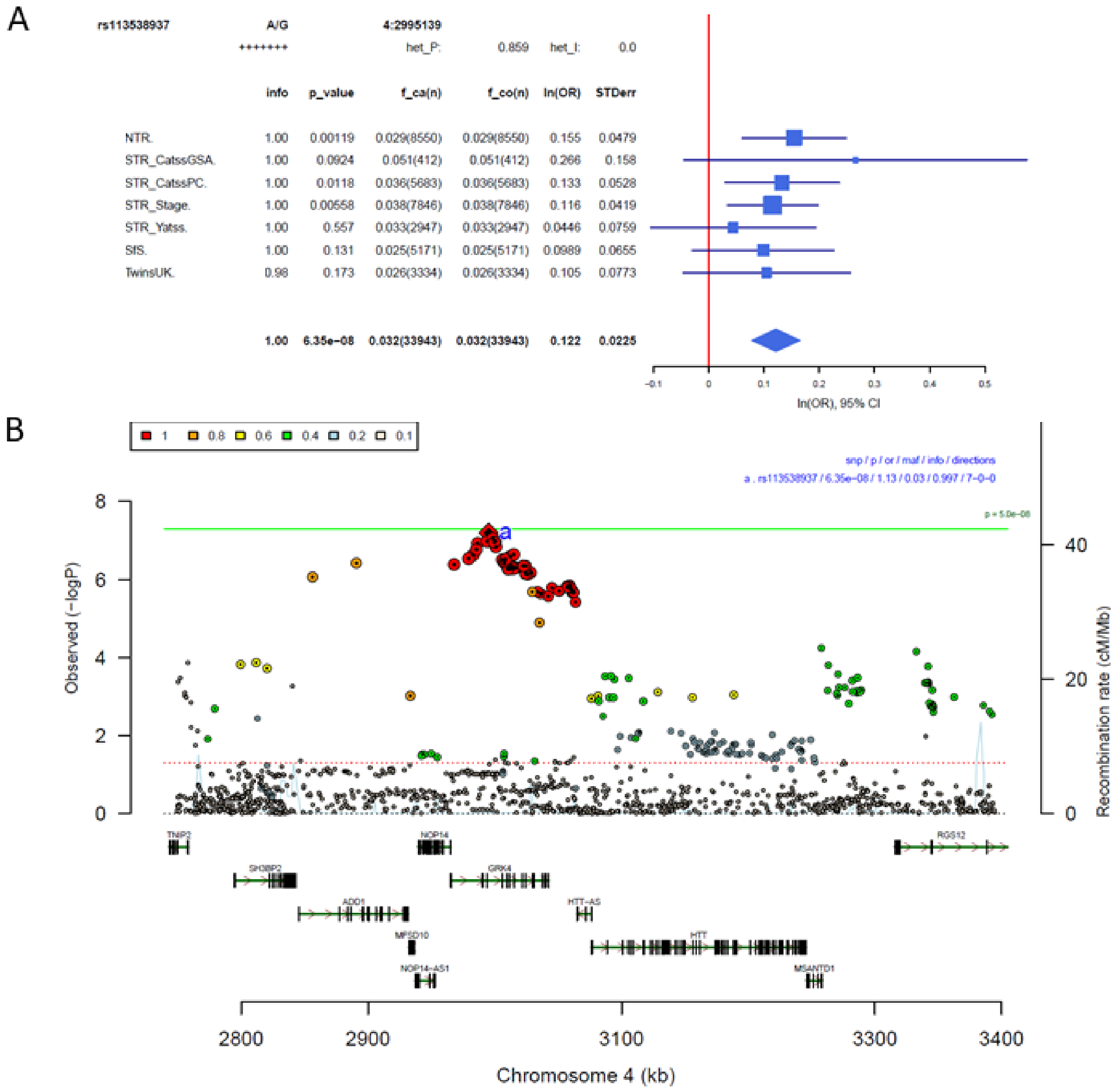

#### 2.2. Compatibility between cohorts

**Supplementary Figure S16:** Manhattan-plot and QQ-plot of heterogeneity test.

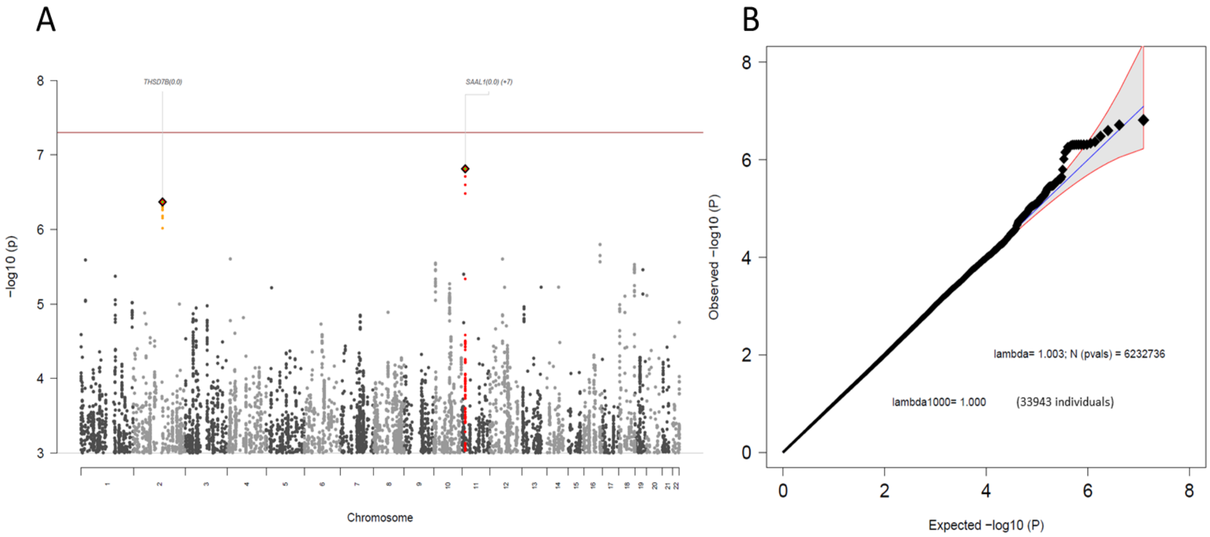

**Supplementary Figure S17:** Manhattan-plot (A) and QQ-plot (B) of the leave-one-out (LOO) GWAS analysis, leaving out STR. N = 17 055.

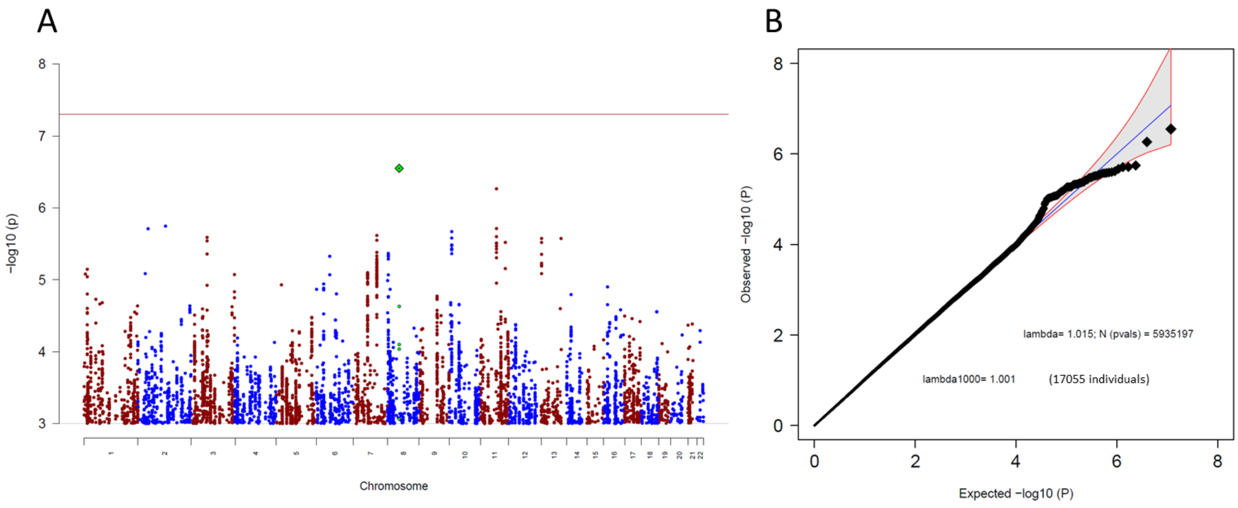


**Supplementary Figure S18:** Manhattan-plot (A) and QQ-plot (B) of the leave-one-out (LOO) GWAS analysis, leaving out NTR. N = 25 393.

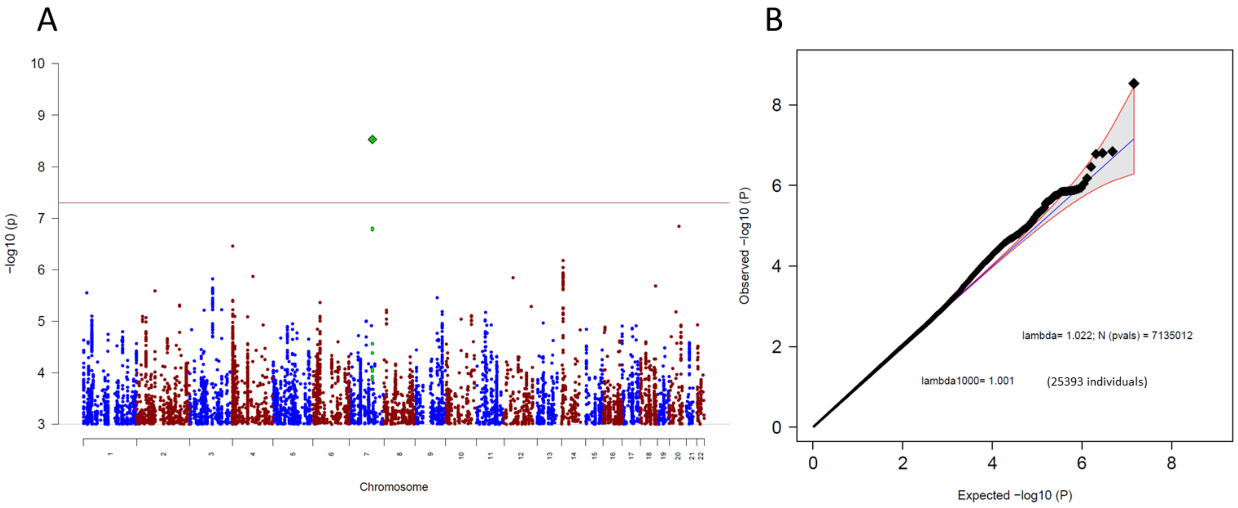

**Supplementary Figure S19:** Manhattan-plot (A) and QQ-plot (B) of the leave-one-out (LOO) GWAS analysis, leaving out SfS. N = 28 772.

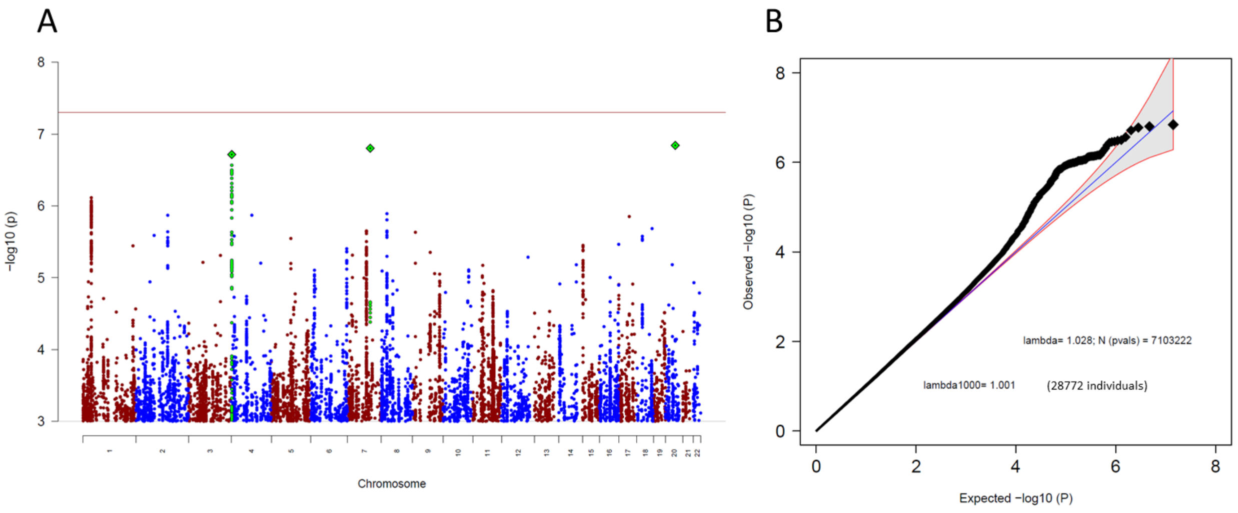


**Supplementary Figure S20:** Manhattan-plot (A) and QQ-plot (B) of the leave-one-out (LOO) GWAS analysis, leaving out TwinsUK. N = 30 609.

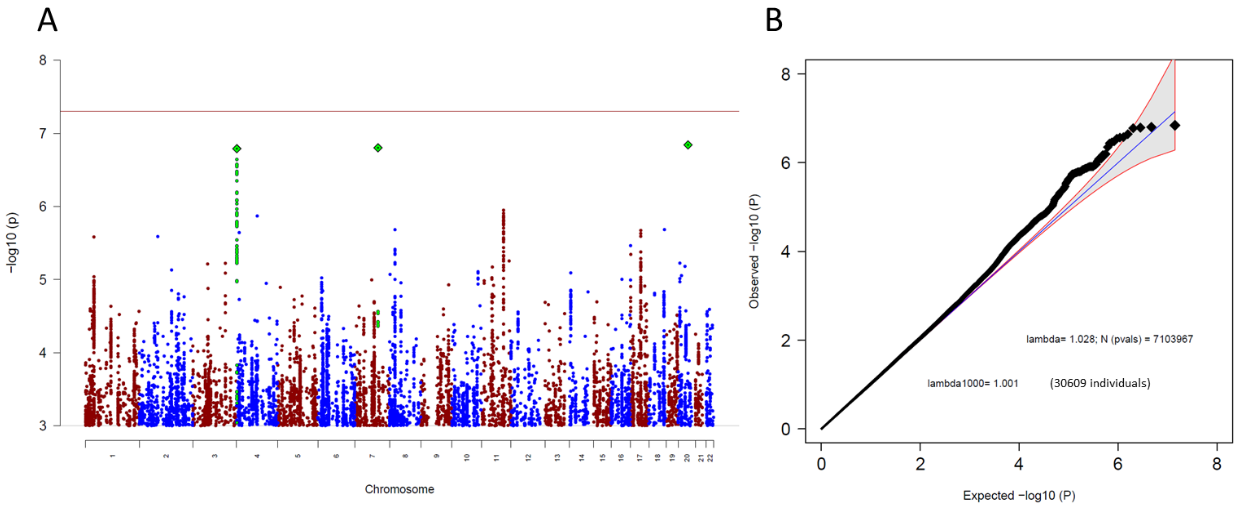

**Supplementary Figure S21:** Genetic correlations (rg) between four LOO OCS datasets and a broad range (N = 97) other phenotypes, assembled into 11 groups (psychiatric, substance, cognition/socioeconomic status (SES), personality, psychological, neurological, autoimmune, cardiovascular, anthropomorphic, fertility, and other). LOO OCS GWAS excluding NTR is presented in blue, excluding SfS is presented in green, excluding STR is presented in yellow, and excluding TwinsUK is presented in pink. Error bars represent 95% confidence intervals; black circles around the dot of estimation indicate significant association after FDR correction for multiple testing. FDR correction was conducted separately for each LOO OCS dataset.

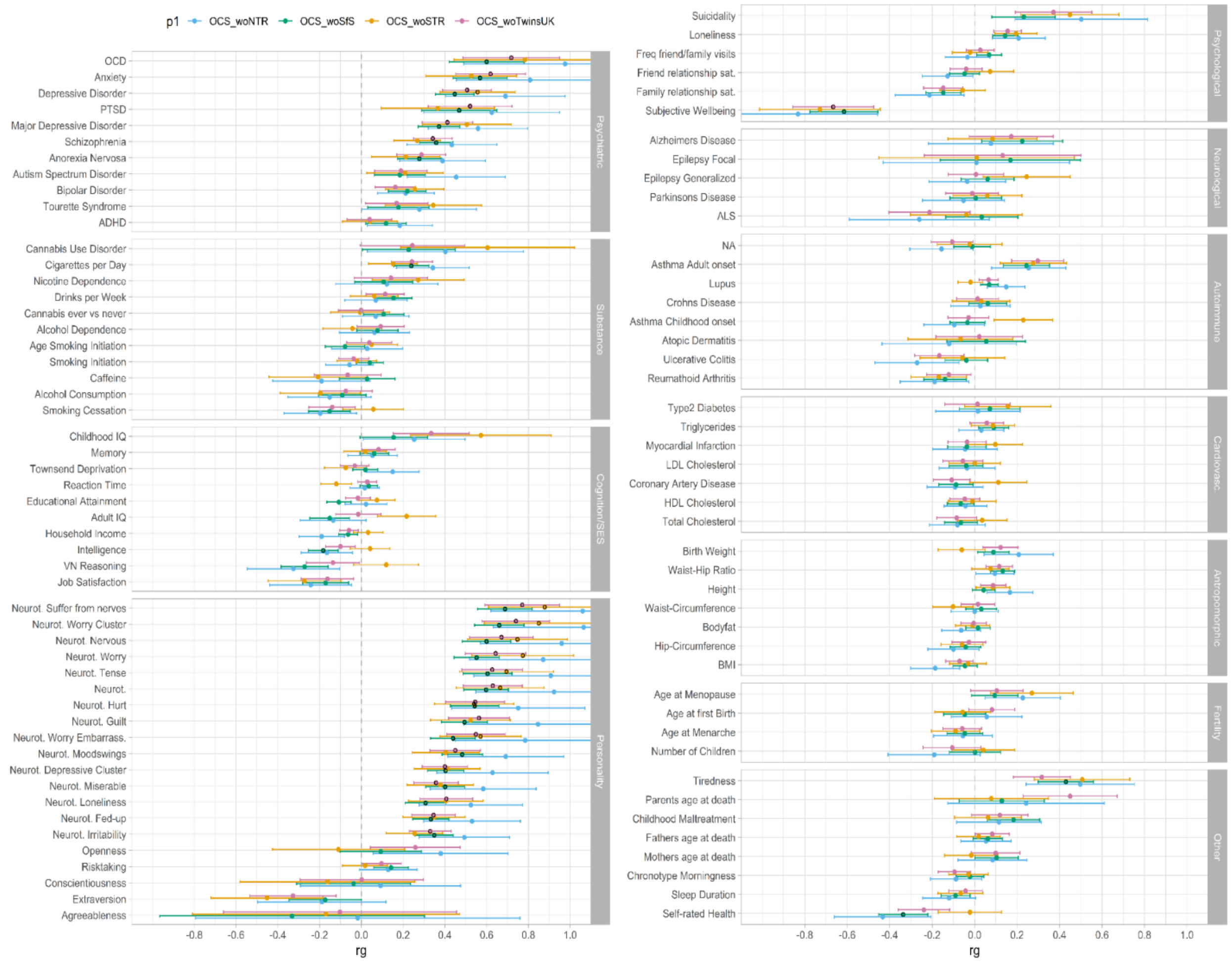
